# The transcriptomic landscape of normal and ineffective erythropoiesis at single cell resolution

**DOI:** 10.1101/2022.08.01.22278133

**Authors:** Raymond T. Doty, Christopher G. Lausted, Adam D. Munday, Zhantao Yang, Xiaowei Yan, Changting Meng, Qiang Tian, Janis L. Abkowitz

## Abstract

Ineffective erythropoiesis, the death of maturing erythroid cells, is a common cause of anemia. To better understand why this occurs, we studied the fates and adaptations of single erythroid marrow cells from individuals with Diamond Blackfan anemia (DBA), del(5q) myelodysplastic syndrome (del(5q) MDS), and normal controls, and defined an unhealthy (vs. healthy) differentiation trajectory, using velocity pseudotime and cell surface protein assessment. The pseudotime trajectories diverge immediately after the cells upregulate transferrin receptor (CD71), import iron, and initiate heme synthesis, although cell death occurs much later. Cells destined to die highly express heme responsive genes, including ribosomal protein and globin genes. In contrast, surviving cells downregulate heme synthesis, while upregulating DNA damage response, hypoxia and HIF1 pathways. Surprisingly, 24±12% of cells from controls follow the unhealthy trajectory, implying that heme also regulates cell fate decisions during normal red cell production. Del(5q) MDS (unlike DBA) results from somatic mutations, so many normal (unmutated) erythroid cells persist. By independently tracking their trajectory, we gained insight into why they cannot expand to prevent anemia. In addition, we show that intron retention is especially prominent during red cell differentiation. The additional information provided by messages with retained introns also allowed us to align data from multiple independent experiments and thus accurately query the transcriptomic changes that occur as single erythroid cells mature.

## INTRODUCTION

Erythropoiesis is a complex process in which bone marrow progenitors multiply and differentiate into mature, enucleated red blood cells. Humans produce 2.3 ×10^6^ red cells each second, and this can increase 5-10 fold when needed, as documented by the reticulocyte count. To allow high and adaptable output while maintaining cell integrity, erythropoiesis needs to be tightly regulated, efficient and “effective”. Should excessive numbers of erythroid precursors die while maturing in the bone marrow, anemia ensues, and erythropoiesis is termed, “ineffective”. Patients with Diamond Blackfan anemia (DBA), thalassemia, and the anemia of myelodysplastic syndromes (MDS)^1^ present with macrocytic anemia due to ineffective erythropoiesis. However, despite previous studies by our group and others,^2-12^ why erythroid cells die while maturing in the marrow is unclear.

The sequential events of effective erythropoiesis, in contrast, are well understood. Hemoglobin comprises 95% of red cell protein content and the coordination of heme and globin is essential. As BFU-E (earliest erythroid progenitors) mature to CFU-E and proerythroblasts, their cell surface transferrin receptor (TfR1, CD71) is upregulated, allowing an influx of iron and upregulation of heme synthesis. Heme (a toxic chelate) next induces the transcription and translation of globin (a protein) by removing inhibitors Bach1 and HRI, respectively. At the time when iron is plentiful and heme synthesis is robust, yet globin is insufficient, CFU-E and early erythroid precursors depend on FLVCR (a cytoplasmic heme export protein) to export unneeded intracellular heme and avoid heme and ROS-mediated damage via apoptosis and ferroptosis (a ROS-dependent process amplified by p53).^13-15^ Later in erythroid differentiation, many cell-intrinsic mechanisms adjust red cell size (MCV) in response to heme or globin availability^16,17^ to assure that the <u>m</u>ean <u>c</u>oncentration of <u>h</u>emoglobin per mature red <u>c</u>ell (MCHC) remains optimal for tissue oxygen delivery and systemic CO2 exchange.

Erythropoiesis is commonly tracked by flow cytometry since maturing erythroid cells sequentially express the cell surface markers CD71, CD36, Glycophorin A and band 3, and these markers correlate well with developmental stage and cell function.^18-20^ Much recent data, however, suggest that differentiation, including erythropoiesis, is a continuum rather than discrete steps.^21-23^ Ostensibly, each cell faces obstacles during maturation and its survival or death depends on its internal programming and its ability to overcome successive challenges or compensate for them, providing a compelling rationale for single cell methods. However, when cells are grouped by flow-cytometric sorting or transcriptional clustering, the observed changes reflect both the insults and the compensations.^12,24,25^

Here, we studied normal (effective) erythroid differentiation and the ineffective erythropoiesis of DBA and del(5q)MDS directly and during in vitro culture. After quantitating the intensity of cell surface markers and total transcriptome of individual cells, we generated a transcriptional pseudotime order with traditional erythroid cell stages. This allowed us to identify distinct developmental trajectories within the normal erythroid differentiation pathway. We defined healthy and unhealthy trajectories. The unhealthy trajectory contained cells with decreased expression of mitochondrial-targeted genes and erythroid genes, yet increased expression of heme-responsive genes. This trajectory diverged right after the upregulation of the transferrin receptor CD71, influx of iron and upregulation of ALAS2, and ultimately led to cell death. 73±2% of DBA and 57±4% of del(5q)MDS cells followed this trajectory, while DBA and del(5q)MDS cells that reduced heme synthesis followed the healthy pseudotime trajectory. Upregulation of hypoxia and HIF1 signaling pathways also selected mutant cells for preferential survival.

Surprisingly, 24±12% erythroid progenitors from normal volunteers followed the unhealthy pseudotime trajectory, and their gene expression patterns during differentiation resembled unhealthy DBA and del(5q)MDS cells. This suggests that balancing heme with globin is a regulatory step that provides a physiological (as well as pathophysiological) way to adjust red cell production.

In addition, our observations help to address an enigma in low risk MDS patients – at presentation, often only 50-75% of cells derive from the mutant clone, yet anemia is prominent. Why residual normal (unmutated) cells cannot expand and compensate is unknown. In the two del(5q)MDS patients at the time of study, 25-35% of their bone marrow cells were normal. We developed a method to independently track the differentiation of erythroid precursors with a chromosome 5q deletion and those with an intact chromosome 5. This allowed us to resolve insult from compensation and provided new insights into the cell-extrinsic regulation of erythropoiesis. We also developed methods to assess intron retention as erythroid differentiation proceeds which allowed us to reliably align cells and directly compare data from separate experiments, a computational approach that should be useful to others.

## Methods

### Study design and data analysis

To investigate erythroid differentiation during normal and ineffective erythropoiesis, lineage-depleted marrow cells were analyzed by flow cytometry and processed for antibody barcoding and single-cell RNA sequencing (scRNAseq) either directly (day 0) or after 3 or 6 days of erythroid expansion culture (Figure 1A, supplemental Figure 1). Marrow samples were collected after written informed consent with a University of Washington IRB approved protocol for this study. We generated 41 scRNAseq samples from two DBA patients, two del(5q)MDS patients, and seven normal controls (Table 1), and sequenced 74,214 individual marrow cells after excluding doublets. Cells containing over 15% mitochondrial reads were considered to be dead and excluded (4869 removed, 69345 retained). Also, genes expressed in less than 20 cells were filtered to assure robust statistical analysis. At least one control sample was included in each experiment and some samples were included in multiple experiments to aid our comparisons of independent experiments and to demultiplex samples (Table 2). Sequence reads were aligned to the human genome (GRCh38) and the results saved to bam files which were analyzed by Velocyto^28^ and transcript splice quantification saved as loom files. Loom files were aggregated by ScanPy.^29^ Transcript counts were normalized (1×10^4^ UMIs per cell), log-transformed, and each gene was scaled to unit variance.

**Figure 1.**
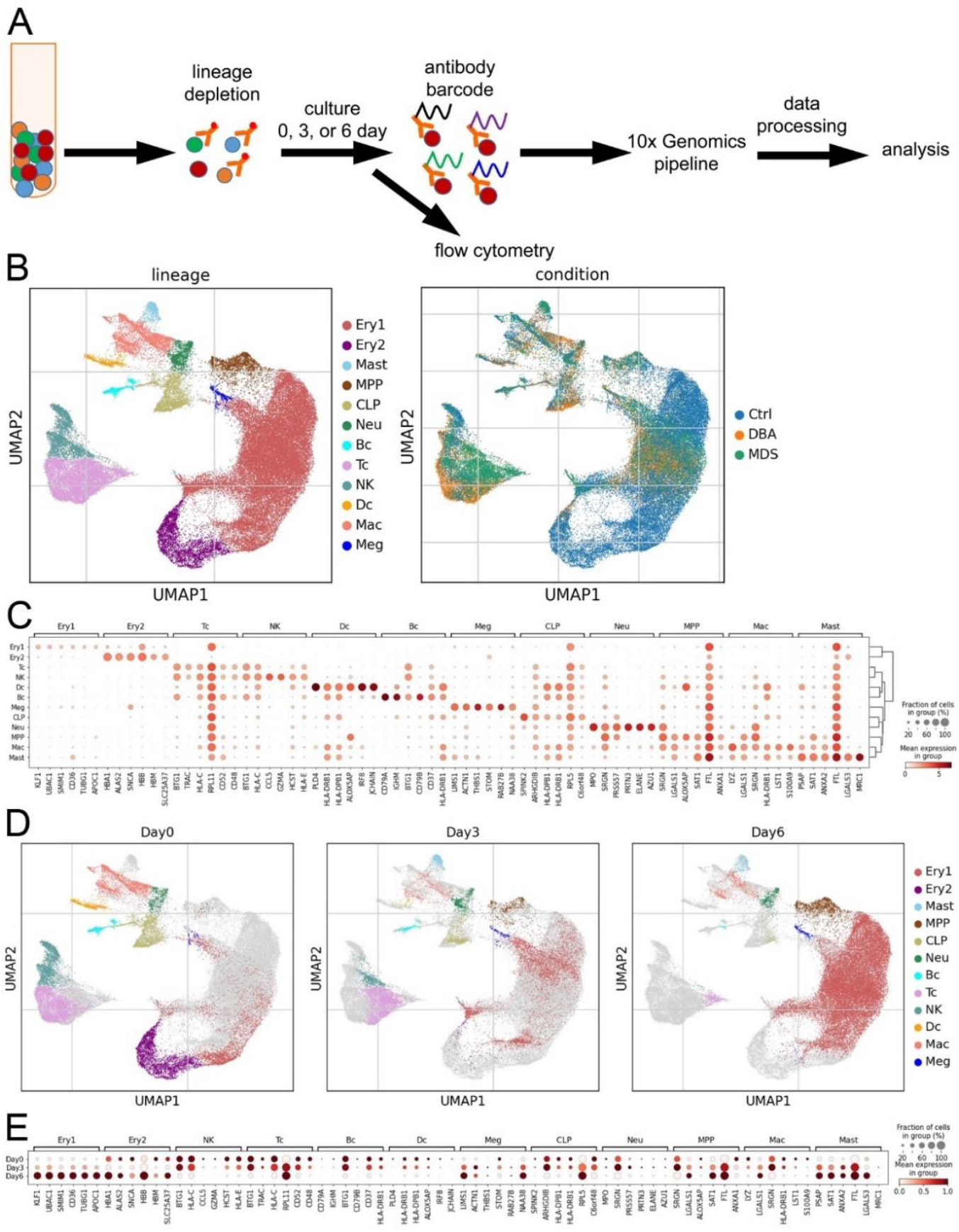
Single-cell transcriptome of erythroid cell enriched bone marrow. (**A**) The experimental design used to enrich erythroid marrow precursors and label surface proteins for quantification during scRNAseq. (**B**) UMAP visualization of all bone marrow mononuclear cells identified by cell lineage and type of sample. (**C**) Dot plot of gene expression level and frequency showing the 6 highest expressed globally distinguishing genes in each cell lineage cluster. (**D**) Individual UMAP visualizations of all bone marrow mononuclear cells identified by cell lineage for cells analyzed after 0, 3, or 6 days of culture. The small number of cells in the latest stages of erythroid differentiation, as identified by their relatively low diversity of transcripts (ie, orthochromatic erythroblasts or reticulocytes) that persisted from day 0 to day 3 were almost completely absent at culture day 6. Thus the 6 day culture primarily contains the early precursors which were present at very low frequency in marrow. Ex vivo marrow predominately consists of terminal stage precursors. (**E**) Dot plot showing the changes in the distinguishing genes expression level and frequency over time in culture for each cell lineage.

**Table 1.** Samples included in this study.

| **<u>Sample</u>** | **<u>Description</u>** | **<u>Notes</u>** |
| --- | --- | --- |
| N1 | 20-24 year old female | Healthy individual |
| N2 | 20-24 year old male | Healthy individual |
| N3 | 50-54 year old female | Healthy individual |
| N4 | 30-34 year old female | Healthy individual |
| N5 | 60-64 year old male | Healthy individual |
| N6 | 45-49 year old female | Healthy individual |
| N7 | 35-39 year old male | Healthy individual |
| D1<br>D1.2 | 20-24 year old female<br>2<sup>nd</sup> sample 3 years later | DBA, RPS19 L18P mutation, steroid non-responsive, transfusion dependent. DBA patient 1 in ref.<sup>3</sup> |
| D2 | 45-49 year old female | DBA, no identified mutation, steroid responsive but not tolerated, transfusion dependent. Index patient in ref.<sup>27</sup> |
| M1 | 70-74 year old male | del(5q) MDS, 5q13-q33 deletion; 74.5% of cells have the 5q deletion via iFISH. No cytogenetic abnormality besides del(5q). |
| M2 | 60-64 year old female | del(5q) MDS, 5q13-q33 deletion; 65% of cells have the 5q deletion via iFISH. No cytogenetic abnormality besides del(5q). Patient MDS1 in ref.<sup>3</sup> |

**Table 2.** Independent experiments included in this study.

| **<u>Experiment</u>** | **<u>Samples</u>** | **<u>Capture days</u>** | **<u>10x kit version</u>** | **<u>Read length</u>** | **<u>Notes</u>** |
| --- | --- | --- | --- | --- | --- |
| 1 | N1, N2, D1, D2 | Ex vivo, day 6 | V2 | 150 bp |  |
| 2 | N2, N3, D1, D2 | Ex vivo, days 3 & 6 | V2 | 150 bp | includes SPARTA* protein data |
| 3 | N4, D1.2 | Ex vivo, days 3 & 6 | V2 | 75 bp | multiplex samples<sup>‡</sup>, includes CITE-seq<sup>†</sup> |
| 4 | N5, M1 | Ex vivo, days 3 & 6 | V3 | 75 bp | multiplex samples, includes CITE-seq |
| 5 | N6, N7, M2 | Ex vivo, days 3 & 6 | V3.1 | 75/100 bp<sup>§</sup> | multiplex samples, includes CITE-seq |

\*SPARTA: Single-cell Protein and RNA Tagged Antibody
<sup>‡</sup>Multiplex samples reduce the cost of the library preparation in direct proportion to the numbers of independent samples combined into a single library preparation. Sequencing costs are not impacted by multiplexing as the same sequencing depth of each sample in the mixture is needed.
<sup>†</sup>CITE-seq: Cellular Indexing of Transcriptomes and Epitopes by Sequencing
<sup>§</sup>Sequenced once each on NextSeq 550: 75 bp; and NovaSeq 6000: 100 bp

We generated pseudotime trajectories using the *tl*.*velocity_pseudotime* function of scVelo,^30^ which quantifies both spliced and unspliced transcripts. Multiplexed samples were demultiplexed based on natural genetic variation without assistance of donor genome references,^31,32^ and then identified by checking for known mutations, attenuated RPS14 gene expression (5q deletion), or sex-specific gene expression. Gene set scores were calculated using the ScanPy^29^ *score_genes* function. Default parameters were used in all calculations unless noted otherwise. Eleven cell-type scores were calculated using panels of marker genes and their expression was evaluated over time in culture (Figure 1B-E, supplemental Figure 2). Scores were also calculated for cell cycle phase using the gene sets and methods of Satija et al.^33^ Cells were clustered by Leiden community detection^34^ using batch-corrected nearest neighbors.^35^ Differentially-expressed genes (DEG) were identified using the Wilcoxon rank-sum test with Benjamini-Hochberg correction (P<0.05), then gene ontology (GO)-term enrichment analysis was performed using the Enrichr method^36^ which calculates the odds ratio based on the deviation from expected rank. Experiments were analyzed both separately, and combined after normalization and scaling (ScanPy functions), using multiple algorithms. Only results that were concordant across data sets and algorithms are reported. We utilized UMAP^26^ visualization as an atlas for all cell types, diffusion maps for 2 dimensional visualization of erythroid differentiation, and pseudotime scatter plots to visualize changes in individual genes or pathways over differentiation. See supplementary Methods for additional details.

Importantly, the transcriptional program of cultured cells reflected that of cells in vivo. By day 6, all cultures contained ample early erythroid precursors (cluster Ery1). Expression of some globin genes and *TFRC* were increased in culture as expected (supplemental Figure 3A).^37,38^ Additionally, principal component analysis showed minimal differences in cultured cells (supplemental Figure 3B). That these were minor and did not persist throughout differentiation justified the combined analyses of cultured cells with uncultured cells.

### Data Sharing Statement

Raw data are available via the Dryad data repository at https://doi.org/10.5061/dryad.573n5tb77.

## RESULTS

### The scVelo pseudotime algorithm optimally orders differentiating cells

We tested several pseudotime methods and determined that the scVelo pseudotime algorithm^30^ predicted a consistent velocity (differentiation) pseudotime across all experiments. This algorithm includes incompletely spliced mRNAs which are abundant and differentially regulated during erythropoiesis.^39,40^ By integrating velocity pseudotime with the upregulation of specific mRNA and cell surface proteins, we identified BFU-E through orthochromatic erythroblasts (stages B-E) within the pseudotime order, using well-established flow cytometry-based criteria^19,41,42^ (Figure 2A-D; supplemental Figure 4A-D). We confirmed the assigned cell stages were consistent with published data, including increased frequency of cells in S phase of cell cycle during the CFU-E stage (Figure 2E-G; supplemental Figure 4E-G)^19,24,25,43,44^ and used this approach to combine and compare the results of the independent experiments. Pseudotime analysis also confirmed upregulation of *TFRC, HBG1* in cultured cells and subtle changes in PC expression (supplemental Figure 3C-D). As anticipated, the number of distinct genes that are expressed during erythropoiesis peaks at the BFU-E stage and then steadily declines (Figure 2H).

**Figure 2.**
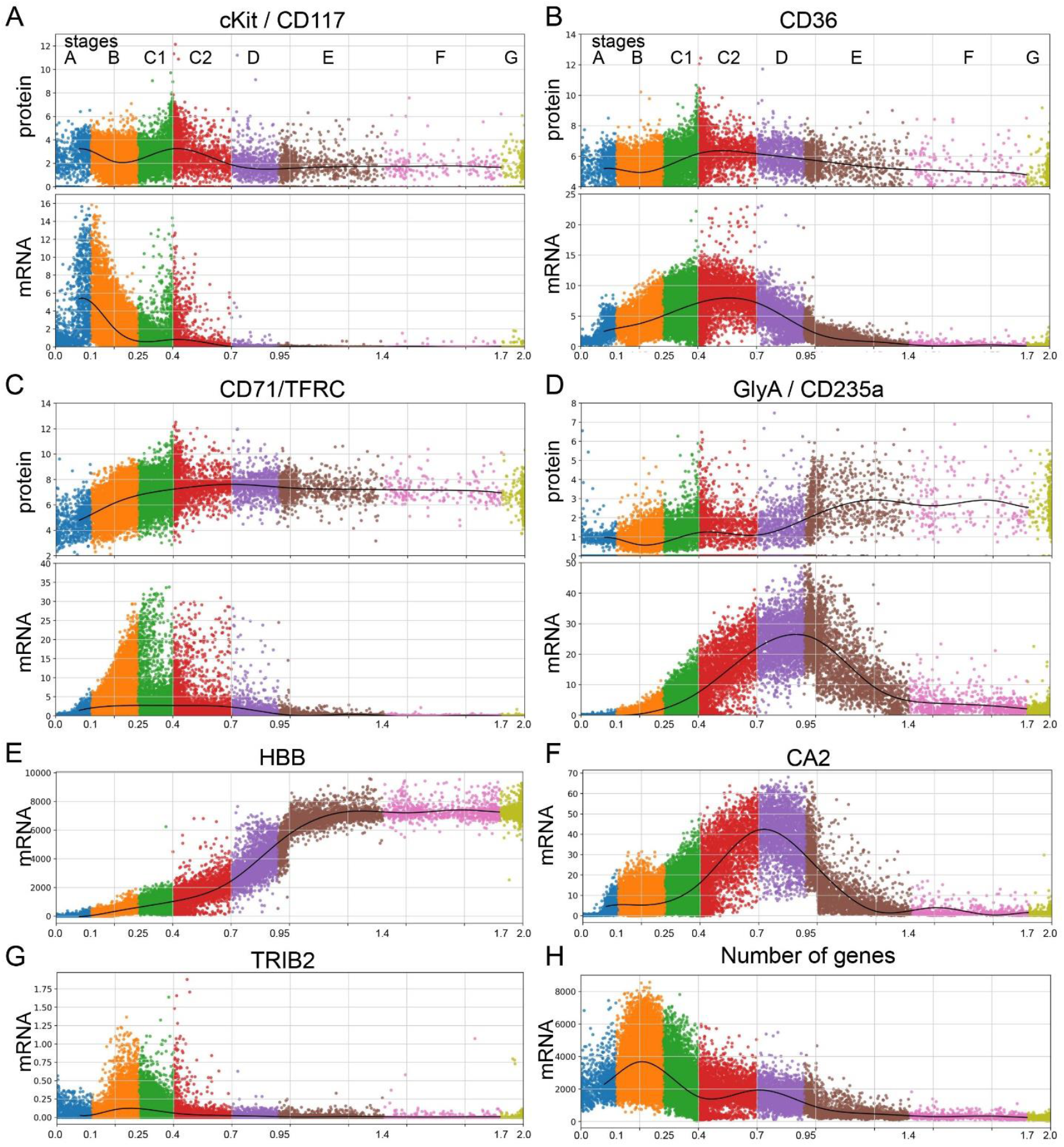
Staging erythroid precursor cells by surface protein marker levels and velocity pseudotime. Velocity pseudotime of erythroid precursor cells, MPP through orthochromatic erythroblasts from Experiments 3-5 showing relative protein and mRNA expression levels of: cKit/CD117 (**A**); CD36 (**B**); CD71/TFRC (**C**); and GlyA/CD235a (**D**) in individual cells. Functional stages of cells have been assigned based upon mRNA and protein expression levels as follows: Stage A (MPP) cells displayed no specific erythroid markers and showed no transcriptional commitment to any lineage. Stage B cells (BFU-E) had high cKit protein and mRNA expression and some CD36 and CD71 protein and mRNA expression. Stage C1 cells (early CFU-E) showed increasing CD36 and CD71 protein but no GlyA protein. Stage C2 (late CFU-E) CD36 mRNA plateaued simultaneously with the upregulation of CD235a/GlyA mRNA expression. Stage D cells (proerythroblasts) had decreasing CD36 expression, increasing GlyA expression, and termination of cKit mRNA and protein expression. Subsequent stages E-G were only present in ex vivo cells from day 0 and largely follow Leiden-based transcriptional clustering. Stage E cells (basophilic erythroblasts) expressed CD36 protein but very low CD36 mRNA expression and high GlyA protein and mRNA expression. Stage F cells (polychromatic erythroblasts) expressed high CD71 and GlyA protein but low GlyA mRNA while stage G (orthochromatic erythroblasts) expressed no CD36 protein but high GlyA protein with no GlyA mRNA. A few reticulocytes (enucleated cells) survived the cryopreservation, but they had low mRNA diversity, extremely high globin transcript levels and were filtered out with high mitochondrial transcripts and low nuclear gene reads (see Methods). (**E**) Log 2 expression of HBB showed steadily increasing expression from BFU-E through basophilic erythroblasts. Velocity pseudotime shows increasing expression of CA2 (**F**) and decreasing TRIB2 (**H**) in individual cells marking the transition from BFU-E to CFU-E on our pseudotime map of erythroid differentiation, consistent with prior studies.^24^ (**F**) Total numbers of genes detected over differentiation shows peak gene diversity at the BFU-E stage followed by decreasing gene diversity throughout terminal erythroid differentiation.

### Intron retention increases as erythroid cells mature

While alternative splicing of mRNA is known to be a key regulatory mechanism,^45-47^ intron retention is also prevalent throughout differentiation. Since unspliced mRNA precedes spliced mRNA, the ratio of unspliced to spliced mRNAs, termed RNA velocity,^30^ can predict developmental trajectories. Interestingly, this ratio increased as erythroid precursors matured to the point that the RNA velocity algorithm predicted a reversed trajectory of differentiation for erythroid cells after the CFU-E stage, but not for any other cell lineages (Figure 3A). Along with the stage-specific changes in intron retention which facilitated velocity pseudotime alignment of diverse experiments, there were gene-specific changes in intron retention. Genes such as *DDX39B, CA2, KLF1*, and *TFRC* showed peak intron retention earlier in differentiation, while intron retention in *GATA1, TAL1*, and globin gene transcripts (e.g., *HBB*) peaked at the terminal stage of erythropoiesis (Figure 3B). We then analyzed the ratio of unspliced to spliced mRNA in bulk ex vivo and cultured marrow via qPCR and observed similar changes (Figure 3C). Thus, erythroid gene expression has broad post transcriptional regulation through intron retention in addition to alternative splicing. As messages with retained introns are not likely to be effectively translated, this provides an additional method to regulate protein expression during erythropoiesis.

**Figure 3.**
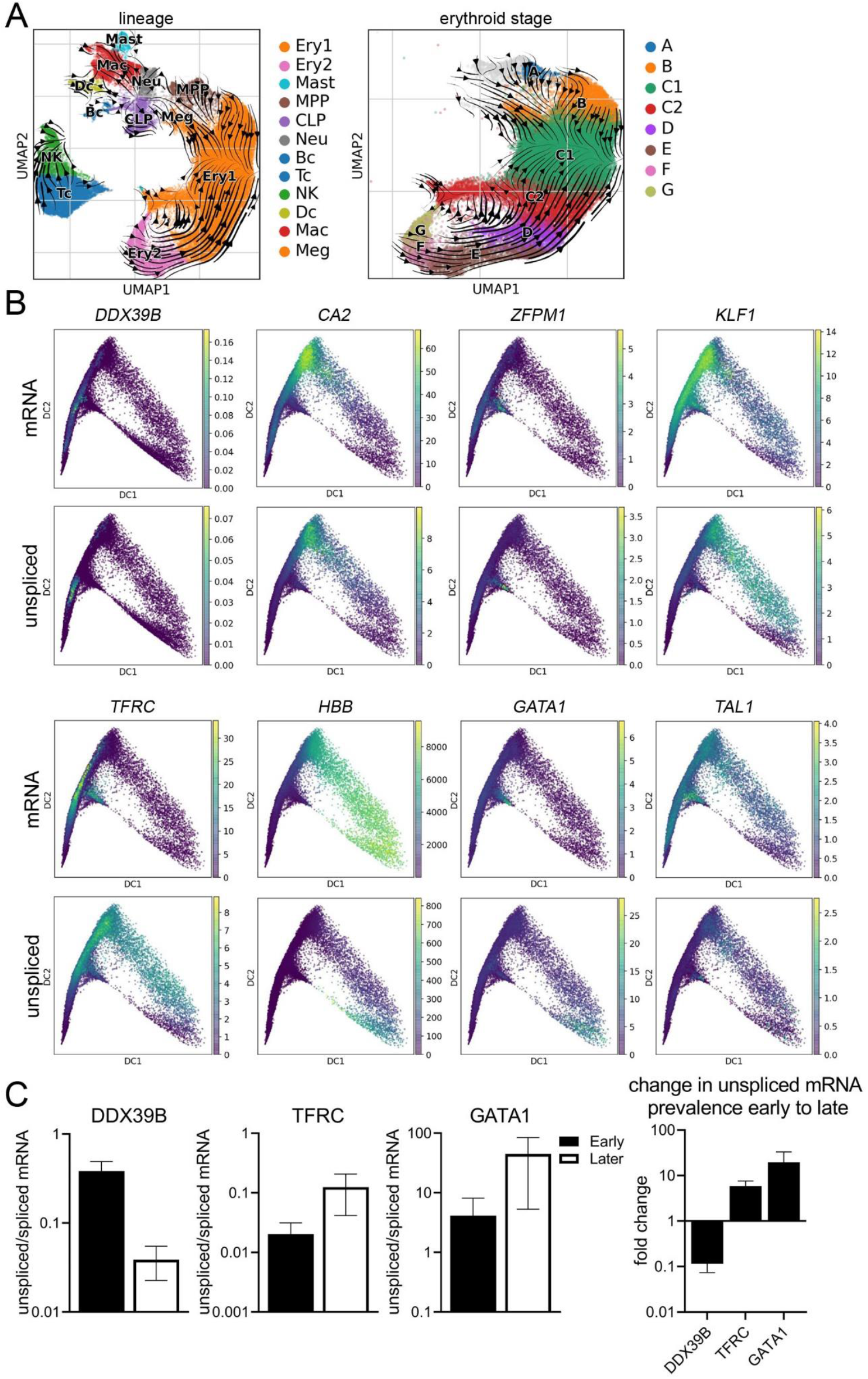
RNA velocity analysis of bone marrow cells shows gene and stage-specific intron retention during terminal erythropoiesis. (**A**) UMAP of bone marrow cells from all samples showing the Velocyto RNA velocity stream direction, indicating the direction of increasing mature RNA content and implying the direction of differentiation, overlaid with all cell lineages (left panel) and erythroid stages (right panel). RNA velocity shows differentiation radiates out from the MPP in all lineages. Once erythroid cells reach stage C (CFU-E) the RNA velocity stream maps show reversed velocity stream arrows indicating that there is increasing un-spliced mRNAs during terminal erythroid differentiation. (**B**) Diffusion map projections of all erythroid precursor cells showing the relative expression levels of mature mRNA and unspliced mRNA for DDX39B, CA2, ZFPM1, KLF1, TFRC, HBB, GATA1, and TAL1 showing stage-specific and gene-specific intron retention. DDX39B has peak intron retention very early while CA2 and ZFPM1 have peak intron retention earlier than KLF1 and TFRC while intron retention in globin gene transcripts (e.g., HBB which are needed in extremely high levels late in differentiation), GATA1, and TAL1 peak only at the most terminal stage of erythropoiesis. (**C**) Bulk analysis of intron retention in early stage erythroid cells (day 6 culture) and later stage erythroid cells from whole marrow (day 0) via qPCR. Total mRNA was analyzed for unspliced and spliced mRNA from subjects N4, N7, & D1. The ratio of unspliced to spliced is presented as mean±SEM from early stage cells (day 6) and predominately later stage cells (day 0 marrow) along with the fold change in prevalence of unspliced mRNA from early to later stages to show the trend over differentiation matches that predicted by scRNAseq analysis. As peak mRNA expression and peak unspliced mRNA levels for each gene occur at distinct stages, the regulation of peak intron retention is independently regulated from peak mRNA, and thus protein, expression.

### Transcriptome analysis uncovers two developmental trajectories, which diverge at the CFU-E stage

Erythroid precursor cells from DBA and del(5q)MDS patients upregulated stress response, apoptosis-related, and mitochondrial metabolism genes (supplemental Figure 5; supplemental Table 1). ADA was also highly upregulated in DBA patient cells as expected.^48,49^

As a more rigorous and informative approach, we analyzed the stage-specific transcriptome of each experiment and day to identify the differences between normal controls and anemia patients that were consistent across all experiments and times. When we visualized the changes in transcriptomes over developmental time, we noted two prominent differentiation trajectories that diverged at the CFU-E stage and were present in all samples (Figure 4A, Table 3). The main trajectory (trajectory A) continued through terminal differentiation, while the alternative trajectory (trajectory B) ended by the proerythroblast stage. Each trajectory included cells from both anemia patients and normal individuals (Figure 4A). Our analysis of published scRNAseq data from a study of marrow CD34^+^ cells isolated from DBA patients and normal individuals by Iskander et al.^12^ also showed these two cell-fate trajectories (supplemental Figure 6). Thus we concluded that it was a common feature of erythropoiesis.

**Figure 4.**
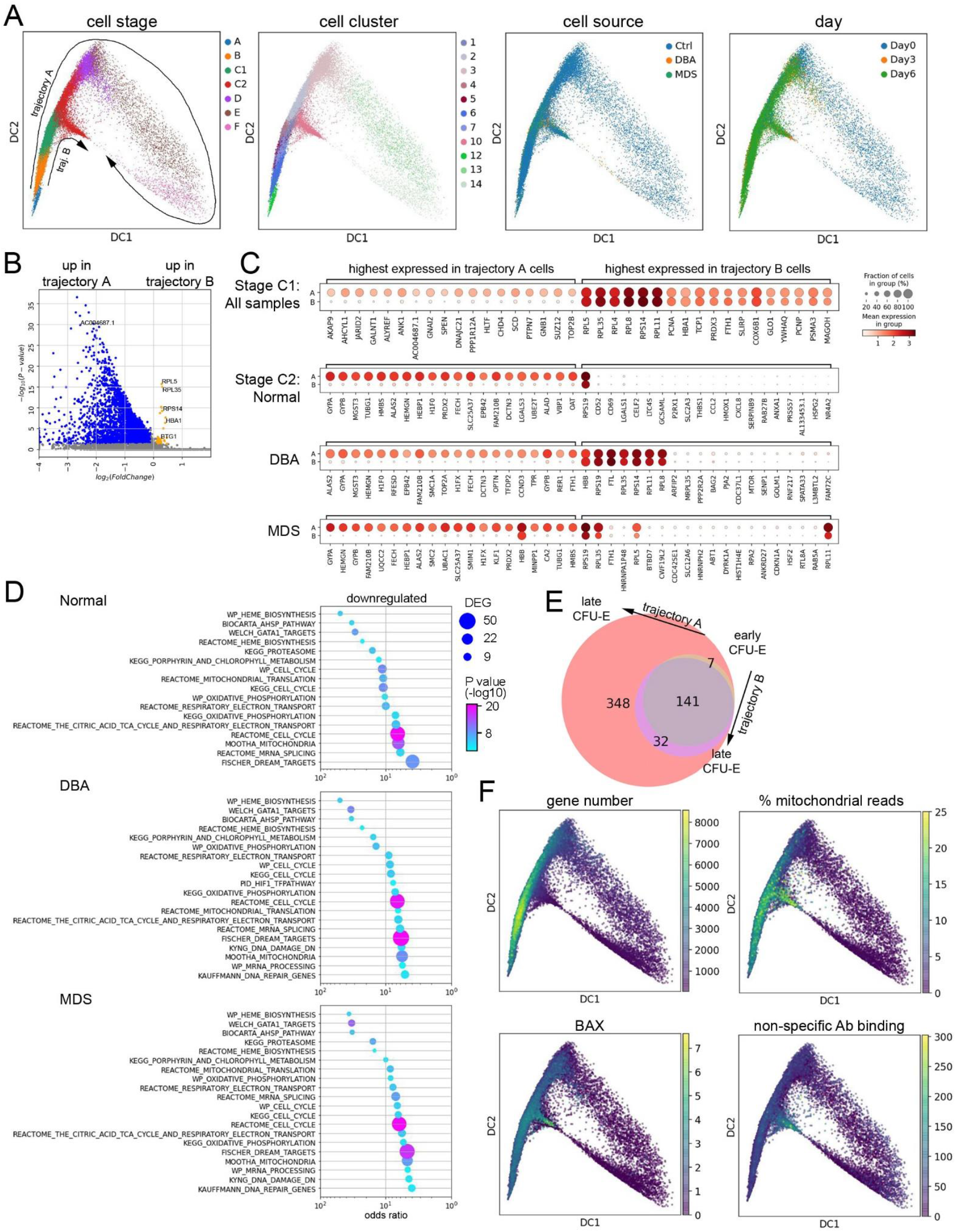
Erythroid gene expression analysis at the single cell level reveals two distinct trajectories. (**A**) Diffusion map visualization of erythroid precursor cells showing MPP and erythroid BFU-E through polychromatic erythroblasts (stages A-F), Leiden cluster identification, cell source, and day of culture. The cell trajectories identified as trajectory B cells from all samples are contained within Leiden cluster 10. Thus trajectory B cells from all samples are transcriptionally similar to each other while being transcriptionally distinct from all trajectory A cells. (**B**) Volcano plot showing significant differences in gene expression in early CFU-E cells (C1 stage) on trajectory B relative to those on trajectory A. See supplemental Table 2 for expression levels of all DEG. (**C**) Dot plots showing relative gene expression and frequency for the top 20 highest expressed genes in early CFU-E cells on trajectory A or B from normal, DBA, and del(5q) MDS samples combined (top, Stage C1), or each sample individually in late CFU-E cells (Stage C2). (**D**) GO analysis showing the most downregulated pathways in late CFU-E stage cells on trajectory B compared to those on trajectory A in normal, DBA, or del(5q) MDS samples. The pathways are sorted by the odds ratio with the number of DEG in the pathway indicated by dot size and the adjusted P value indicated by dot color. See supplemental Table 2 for complete lists of DEG. (**E**) Venn diagram showing the numbers of newly expressed genes (>100 copies/cell) as cells progress from early CFU-E (C1 stage) to late CFU-E (C2 stage) on either trajectory A or trajectory B. (**F**) Diffusion map visualization of erythroid precursor cells (BFU-E through orthochromatic erythroblasts) showing total gene expression numbers, the percent mitochondrial reads, *BAX* expression levels, and non-specific CITE-seq antibody binding quantification.

**Table 3.**
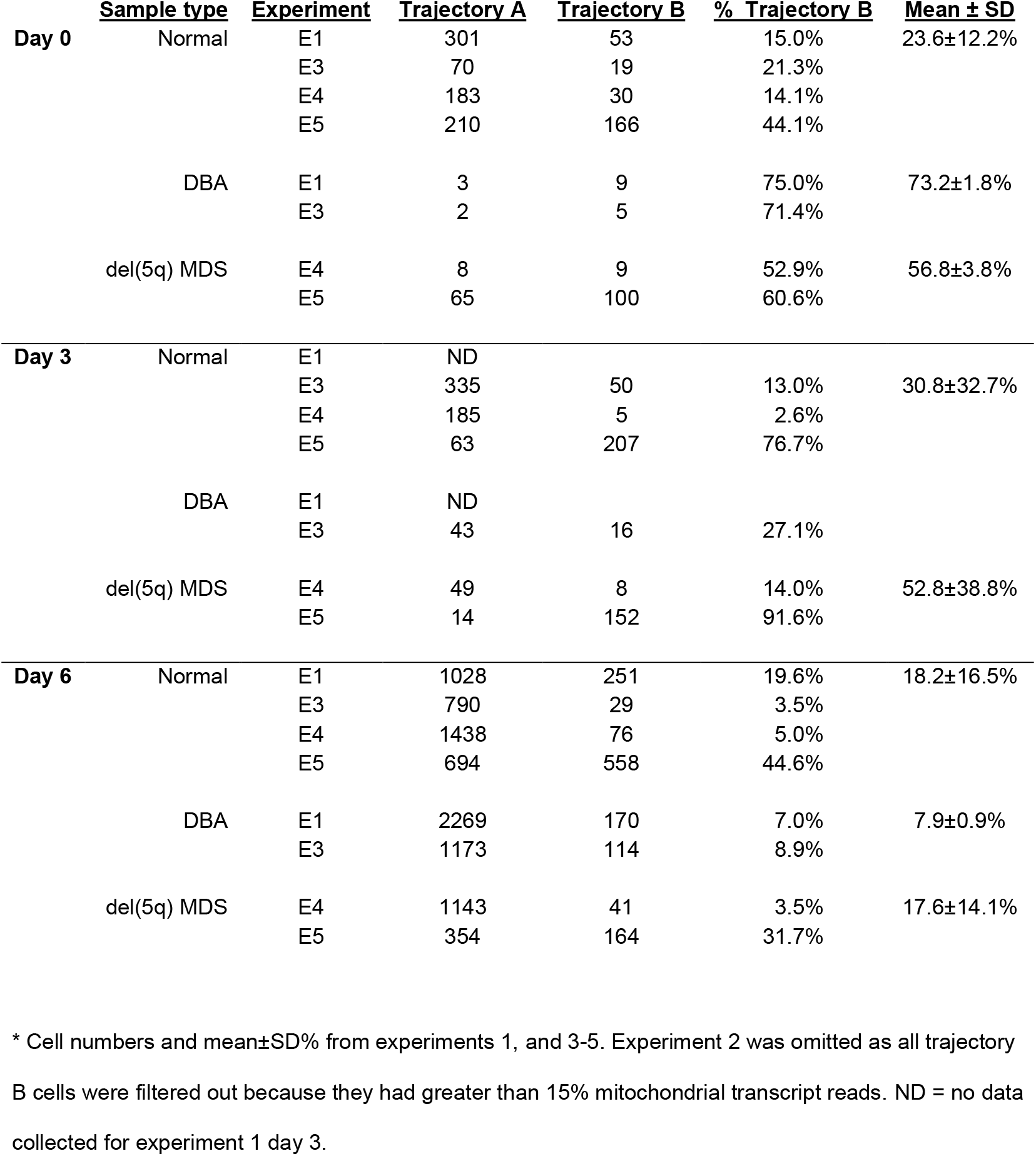
Frequency of BFU-E and CFU-E (stages B & C) cells* on trajectories A and B

### Trajectory B is a death pathway

To better characterize trajectories A vs B, we analyzed gene and pathway expression. The top 20 genes upregulated in trajectory A cells were mostly erythroid differentiation genes and included some cell cycle and mitochondrial oxidative metabolism genes. These genes were poorly expressed in trajectory B cells (Figure 4B-D; supplemental Table 2). While many new genes were upregulated when cells progressed from early CFU-E (C1) to late CFU-E (C2) along trajectory A,10-fold fewer new genes were expressed when cells progressed from early CFU-E to late CFU-E along trajectory B (32 vs 348, Figure 4E). GO enrichment analysis replicated this. Most pathways were down-regulated in late CFU-E on trajectory B from normal, DBA, and del(5q)MDS patient samples (Figure 4D; supplemental Table 2), indicating failed differentiation.

24±12% of day 0 (uncultured) B&C stage cells from normal individuals followed trajectory B while 2-3-fold more day 0 cells from DBA and del(5q)MDS patients (73±2 and 57±4 %, respectively) followed trajectory B (Table 3, also see supplementary Figure 6C-D). However, culture allowed more patient cells to follow trajectory A and complete erythroid differentiation. This finding confirms our previous work showing that culture enables erythroid precursors from DBA and del(5q)MDS patients to survive.^3^

Protein barcoding and pseudotime analysis placed the trajectory branch point during the CFU-E stage, thus after iron import via CD71 and when heme synthesis is first upregulated (Figures 2, 5A-B). Despite significant transcriptional differences between cells on trajectory A versus trajectory B (Figure 4B-D), the cell surface expression of CD36, CD71, and GlyA on cells as they progressed along both trajectories were sufficiently similar to prevent prospective flow-based resolution of these trajectories (Figure 5B).

**Figure 5.**
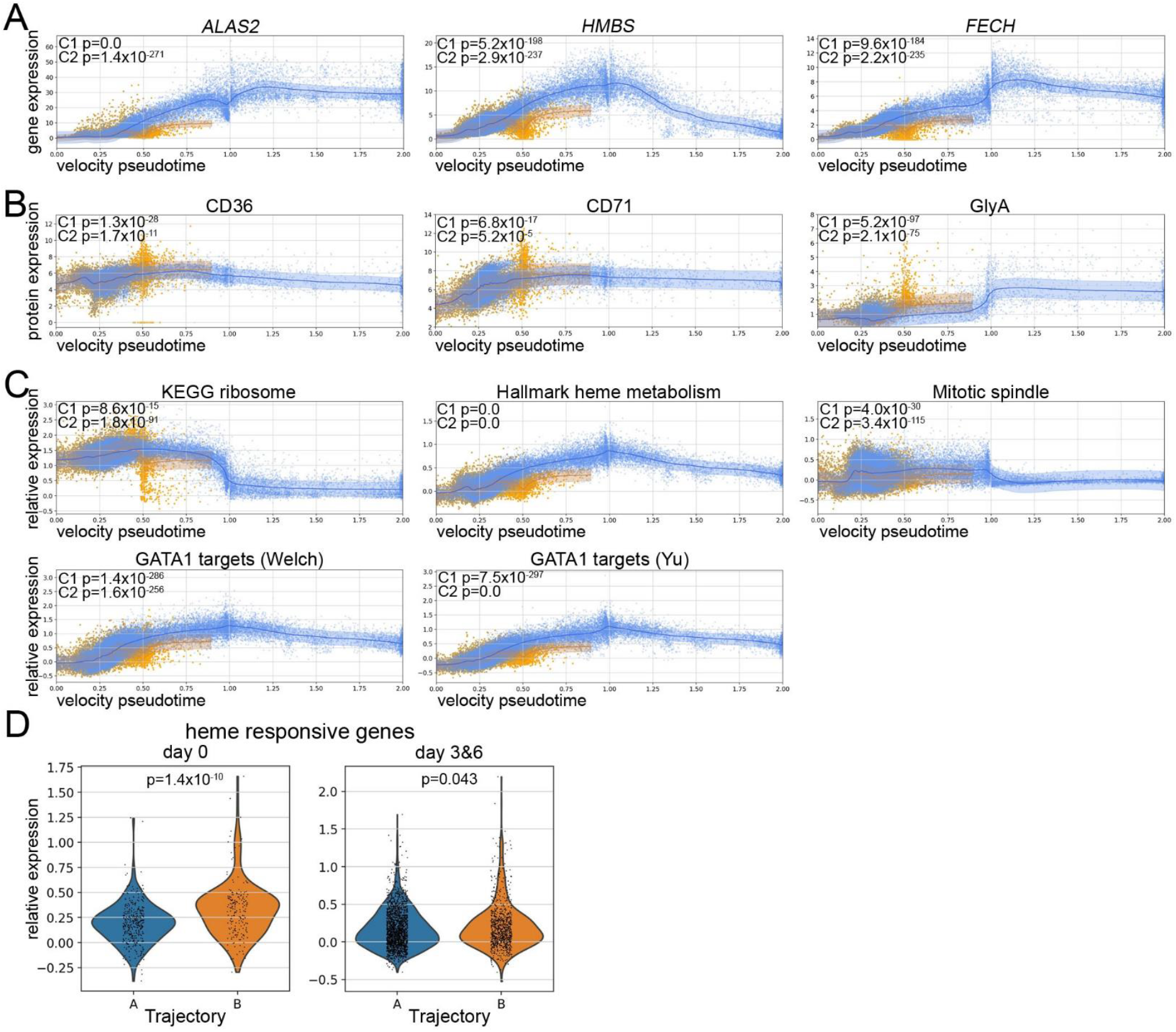
Trajectory B cells are identified by changes in transcription but not cell surface protein expression. Velocity pseudotime map of all cells from normal volunteers, DBA patients, and del(5q) MDS patients mapped to trajectory A (blue) or trajectory B (orange) showing expression levels of heme biosynthetic genes (**A**) ALAS2, HMBS, and FECH are decreased in cells on trajectory B while (**B**) CD36, CD71, and GlyA proteins are not decreased, but slightly increased, in trajectory B cells. (**C**) Velocity pseudotime of all cells showing expression of genes in the ribosome, hallmark heme metabolism, mitotic spindle, and GATA1 targets^63,64^ pathways. While ribosome genes are initially upregulated in early CFU-E on trajectory B, the other pathways are downregulated in both early and late CFU-E cells on trajectory B. The mean expression levels are indicated by the trend line with a 1 SD interval shaded. (**D**) Violin plots showing upregulated expression of heme responsive genes from Liao et al.^50^ in late CFU-E (stage C2) cells on trajectory B at day 0 or after culture (Day 3&6). The upregulation is less pronounced in cultured cells suggesting that either heme responsive gene expression, or heme synthesis itself, is attenuated in culture.

As a further demonstration that trajectory B ultimately leads to cell death, trajectory B cells showed a rapid decrease in nuclear gene expression and a rapid increase in the percent of mitochondrial reads (Figure 4F), indicating a loss of membrane integrity. Additionally, these cells had the highest expression of BAX and high levels of non-specific CITE-seq antibody binding. A cell’s decision to follow trajectory B (per transcriptomics) occurs well before its ultimate demise (Figure 5A-B). This implies that there is time during early erythroid differentiation when therapeutic interventions could amend cell fate.

### Transcriptional analyses implicate heme excess in cell death

While very few genes were upregulated in trajectory B compared to trajectory A cells, the highest expressed genes in trajectory B cells in all samples were ribosomal protein genes. These heme-responsive genes were significantly upregulated in early CFU-E, the stage where trajectories A and B initially diverge (Figure 4C & 5C). We also noted a significant increase in heme-responsive gene expression, as defined in Liao et al.,^50^ in late CFU-E (C2 stage) trajectory B cells (Figure 5D), the stage where heme synthesis intensifies. In addition, pseudotime analysis revealed significant downregulation of GATA1 targets, hallmark heme metabolism, mitotic spindle, and ribosome pathway genes in trajectory B versus trajectory A cells (Figure 5C). This is in addition to the downregulated mitochondrial electron transport and mRNA splicing genes revealed by GO analysis (Figure 4D). As these findings were present in trajectory B cells in all samples and are similar to the transcriptional alterations seen in our single cell analyses of *Flvcr*-deleted and Rpl11 haploinsufficient mice,^51,52^ the data implicate heme excess in cell demise. The transcriptional alterations in trajectory B cells are also similar to those seen in in vitro studies of normal human CD36^+^ erythroid cells 15 minutes after heme synthesis is induced by adding ALA (aminolevulinic acid).^51^ That the transcriptomes of the erythroid cells from healthy individuals on trajectory B resembled the transcriptomes of DBA and del(5q)MDS erythroid cells on trajectory B was an unexpected finding, and suggests that heme regulates normal, as well as pathological, erythroid maturation.

### DBA and del(5q)MDS cells attempt to compensate for excess heme

Although all cells are impacted, some DBA and del(5q)MDS cells can follow trajectory A and survive. To determine how this occurs, we performed GO analyses of erythroid precursor cells at each stage of trajectory A differentiation and compared this to the samples from healthy individuals (Figure 6; supplemental Tables 3-4). This allowed us to identify alterations that were common to both DBA and del(5q)MDS as differentiation occurs.

**Figure 6.**
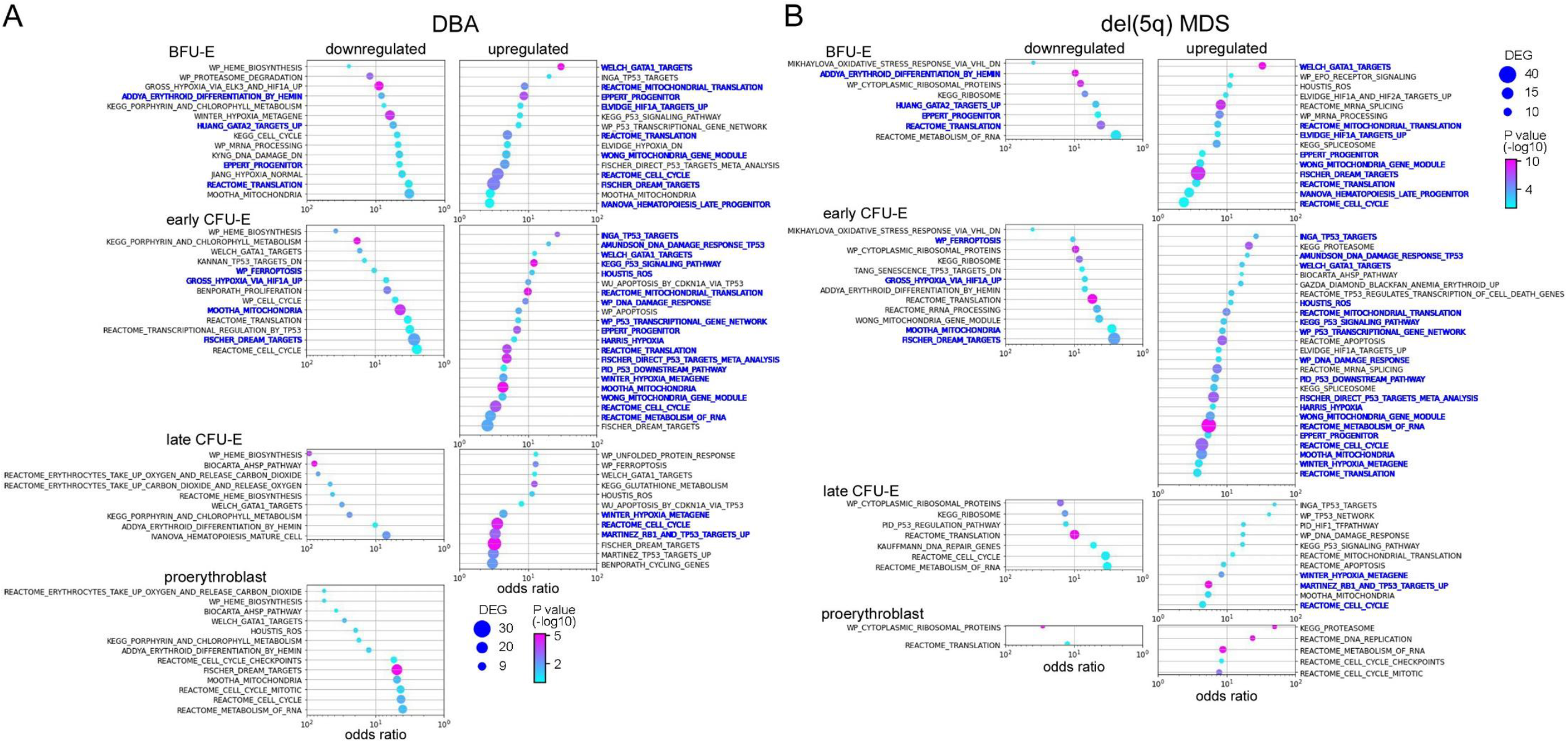
Erythroid cells from DBA and del(5q) MDS patients share many transcriptional alterations. GO analysis of erythroid precursor cells on trajectory A comparing cells from (**A**) DBA patients and normal individuals and (**B**) del(5q) MDS patients and normal individuals showing upregulated and downregulated pathways. Pathways with the same stage-specific alterations in both DBA and del(5q) MDS cells are labeled in blue. The pathways are sorted by the odds ratio with the number of DEG in the pathway indicated by dot size and the adjusted P value indicated by dot color. See supplemental Tables 3-4 for the DEG lists.

Cell cycle pathways (cell cycle, cell cycle mitotic, dream targets pathways) were significantly dysregulated in DBA cells on trajectory A when compared to normal cells on trajectory A, indicating that DBA trajectory A cells were stressed. Ribosomal protein genes (reactome translation pathways) were upregulated in early DBA trajectory A erythroid cells (stage A-C1; Figure 6A; supplemental Table 3), consistent with elevated heme levels.^51,52^ p53 signaling pathway, p53 targets, and hypoxia pathway genes were upregulated later. Notably, DBA trajectory A erythroid cells had significantly reduced expression of heme biosynthesis genes and related pathways (heme biosynthesis, porphyrin and chlorophyll metabolism, and alpha-hemoglobin stabilizing enzyme (AHSP) pathways) beginning at the BFU-E stage. While GATA1 was transcriptionally upregulated, GATA1 targets were not uniformly upregulated (heme biosynthetic gene targets were down regulated while other targets such as globin genes were upregulated). Thus, surviving DBA cells (trajectory A) downregulate heme production and upregulate globin transcription. This offsets their slowed protein (globin) translation from ribosomal haploinsufficiency, mitigates heme toxicities, and permits survival.

Unbiased GO analysis of erythroid precursor cells on trajectory A from the del(5q)MDS patients showed similar alterations and only minor differences (Figure 6B; supplemental Table 4), indicating a shared pathophysiology with DBA and shared compensatory responses. Interestingly only RPS19 was upregulated in early trajectory A CFU-E del(5q)MDS cells but not other ribosomal proteins. The upregulation of HIF1A targets, hypoxia and DNA damage response genes in trajectory A DBA and del(5q)MDS cells would counteract the ROS produced by excess heme and further promote cell survival. These changes were not seen in trajectory B cells.

The ribosomal haploinsufficiencies present in DBA and del(5q)MDS are sometimes associated with increased p53 activity,^4,53-55^ and the TP53 network and p53 signaling pathway and DNA damage response pathway were commonly upregulated in CFU-E cells on trajectory A as anticipated (Figure 6A-B; supplemental Tables 3-4). Heme-related genes, but not p53 pathway genes, were upregulated in trajectory B cells, including normal controls (see Figure 4B-C). Also, of note, p53 activity is not increased in *Flvcr1*-deleted mice^51^ where excessive intracellular heme results from failed heme export. Together, this indicates that excess heme, and not p53-dependent mechanisms, triggers the demise of trajectory B cells.

### Deconvolution analysis of del(5q)MDS samples shows that both cells containing the chromosome 5q deletion and cells with an intact 5q are abnormal

DBA is an inherited, germline disease and as such, all DBA cells contain the mutation resulting in ribosomal protein haploinsufficiency. However, del(5q)MDS is an acquired clonal disease that originates in a hematopoietic stem/progenitor cell and results in its deletion of chromosome 5q and haploinsufficiency of RPS14. One pathophysiological dilemma is that at presentation del(5q)MDS patients have anemia, even when many unmutated normal hematopoietic stem/progenitor cells remain. At the time of sample collection, 74.5% of marrow cells in the del(5q)MDS patient (M1) had a chromosome 5q deletion, when assayed by iFISH. Although 25.5% of his marrow cells had an intact chromosome 5, he had a profound macrocytic anemia (Hct=20.9%, Hgb=7.0 g/dL, MCV=117 fL; normal ranges 38.3-48.6%, 13.2-16.6 g/dL, and 80-100 fL). We calculated that 77% of erythroid cells contained the 5q deletion using gene expression analysis (Figure 7A-B, supplemental Figure 7).^56^ Studies of the second del(5q)MDS patient were similar as 65% of marrow cells (by iFISH) and 63% of the erythroid cells (by expression) had 5q deleted (Figure 7A-B, supplemental Figure 7).

**Figure 7.**
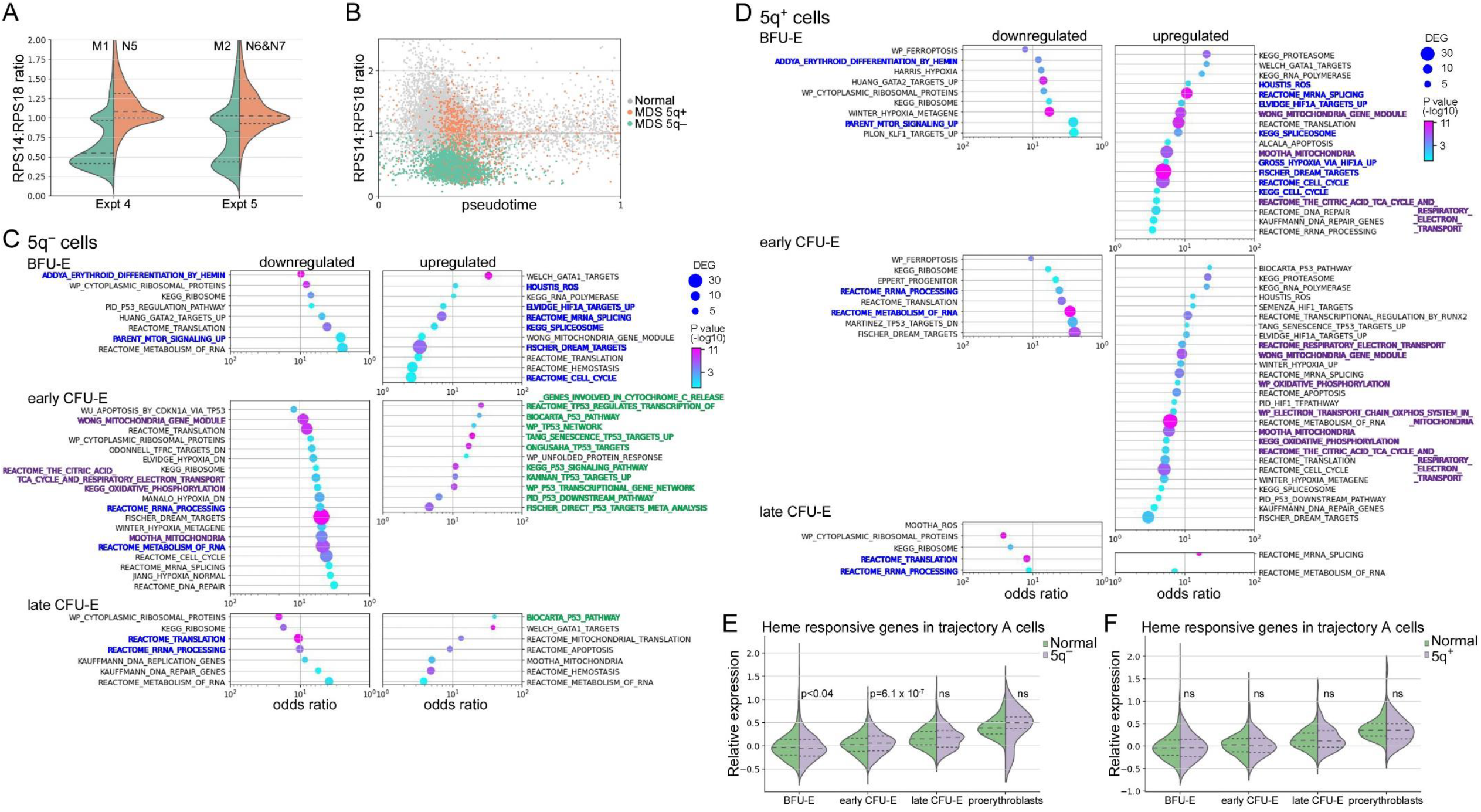
MDS patient cells that have intact chromosome 5q share heme-dependent transcriptional changes that are present in cells that have the 5q deletion. (**A**) Violin plots of the RPS14:RPS18 expression ratio of marrow cells from experiments 4 & 5. (**B**) Velocity pseudotime plots of erythroid cells from experiments 5 & 6 showing the RPS14:RPS18 expression ratio of MDS cells identified as 5q^+^ or 5q^−^ according to the methods of Patel^56^ which evaluates expression of all genes in the 5q13-33 deletion interval. The RPS14:RPS18 expression ratio identified 77% of erythroid cells from M1 and 63% of erythroid cells from M2 were haploinsufficient for RPS14. This was 91% concurrent with reduced gene expression across the 5q13-33 deletion interval, and consistent with iFISH results of 75% and 65% of all marrow cells from the del(5q) MDS patient M1 and M2 respectively (see supplemental Methods and supplemental Figure 7 for additional details). GO analysis of del(5q) MDS patients BFU-E and CFU-E cells on trajectory A that (**C**) have the 5q deletion (5q^−^) or (**D**) have an intact chromosome 5q (5q^+^) to those from normal individuals showing the most upregulated and downregulated pathways. Pathways with similar alterations in both 5q^+^ and 5q^−^ cells are in blue. Pathways related to p53 activation upregulated in 5q^−^ cells are in green while pathways related to mitochondrial oxidative phosphorylation are in purple. The pathways are sorted by the odds ratio with the number of DEG in the pathway indicated by dot size and the adjusted P value indicated by dot color. See supplemental Tables 5&6 for the complete DEG lists. Expression of heme responsive genes in pathway A cells comparing (**E**) 5q^−^ cells to cells from healthy individuals and (**F**) 5q^+^ cells to cells from healthy individuals.

Using this approach we separately analyzed the transcriptomes of the cells on trajectory A with the 5q deletion (5q^−^) and cells with an intact 5q (5q^+^) chromosome (Figure 7C-D). The transcriptomes of the 5q^+^ cells were not normal, but rather showed changes also present in the 5q^−^ cells (and DBA samples). As examples, both the 5q^+^ and 5q^−^ cells showed early upregulation of HIF1A targets, ROS response, cell cycle regulation and spliceosome pathways and down regulation of erythroid differentiation and mTOR signaling pathways (Figure 7C-D; supplemental Tables 5-6). The 5q^+^ cells uniquely upregulated oxidative phosphorylation-related pathways which were downregulated in the 5q^−^ cells. Conversely, the 5q^−^ cells upregulated several p53 associated pathways while the 5q^+^ cells only upregulated the p53 related genes CDKN1A and BAX. Their upregulation in the 5q^+^ early CFU-E cells, suggests p53 activation might contribute to the failure of the 5q^+^ cells to expand and complete erythroid differentiation. When we looked specifically at the expression of heme regulated genes,^50^ we found the 5q^−^ cells on trajectory A, and both 5q^−^ and 5q^+^ cells on trajectory B, had significantly elevated expression of heme responsive genes at the early CFU-E stage (Figure 7E). While trajectory A 5q^+^ cells were able to compensate sufficiently that heme responsive genes were not globally increased (Figure 7F), there was still elevated heme in these cells indicated by the upregulation of oxidative phosphorylation pathways^52,57^ (Figure 7D).

## DISCUSSION

When CFU-E transition to proerythroblast, iron is plentiful and heme synthesis is robust, yet globin is insufficient, and cells depend on FLVCR to export heme and avoid heme and ROS-mediated damage.^3,13,58,59^ When FLVCR is conditionally-deleted in mice, the animals develop a severe macrocytic anemia characterized by excess heme in erythroblasts, excess cytoplasmic ROS, and cell death via apoptosis and ferroptosis.^13-15,59^ Previously, we,^3^ and others^54,60,61^ hypothesized that the macrocytic anemia of DBA and del(5q)MDS, disorders associated with germline or somatic ribosomal protein haploinsufficiencies, respectively, have a related pathogenesis. Here, using a combined dataset to minimize the impact of individual variability, we explored the common pathophysiology of these disorders with single cell analyses. The single cell pseudotime analyses allowed us to dissect the decision-making in an individual cell at designated points along its differentiation timeline and derive novel insights into normal (effective) and failed (ineffective) erythroid differentiation.

Uncultured cells (day 0) from DBA and del(5q)MDS patients and cells after 3 and 6 days of culture distributed onto two trajectories, a healthy one (trajectory A), resulting in effective erythroid differentiation, and an unhealthy one (trajectory B), resulting in ineffective erythroid differentiation and cell death. Data analyses (Figures 5-6) suggest heme excess drives a cell towards pathway B and its ultimate demise. Interestingly, some (24±12%, range 14-44%) erythroid marrow cells from normal controls also followed trajectory B and had gene expression patterns just like trajectory B patient cells (Table 3; Figure 4; supplemental Table 2), suggesting that heme is a broad regulator of effective versus ineffective erythropoiesis. The reason why some normal cells follow pathway B is uncertain, but potentially this assures a ready supply of early progenitors so that erythropoiesis can rapidly expand at times of physiological need.

By the late CFU-E (C2) stage the cells on trajectory B have significant upregulation of heme-regulated genes (Figure 5D) and failed to upregulate erythroid differentiation genes, indicating that one primary cause of death is excess heme. There is substantial time between when cells begin to follow trajectory B and when they die, providing a window for therapeutic intervention, for example with drugs that diminish heme synthesis or chelate iron.

Cells from DBA and del(5q)MDS patients that follow the healthy trajectory A are not normal. Rather they have successfully compensated to avoid fatal damage. First, they have reduced erythroid differentiation genes (including heme biosynthesis genes) which is a direct response to excess heme.^51,52^ Second, they express increased DNA damage response genes, likely to facilitate repair of ROS-mediated cell damage. And third, they have upregulated HIF1A targets and hypoxia response genes, which may facilitate cell survival in light of the reduced heme synthesis. Many of these findings have been previously validated both in vivo and in vitro.^3,51,52^ That culture stabilizes precarious DBA and del(5q) cells allowing a greater number to survive and effectively differentiate (follow trajectory A) suggests that factors present in culture help cells compensate. That some patients with DBA spontaneously remit^62^ implies that cell-extrinsic factors can also improve the in vivo differentiation of erythroid cells haploinsufficient for a ribosomal protein. It seems possible that drugs which mimic these compensations might ameliorate DBA or del(5q)MDS anemia.

We also found that the cells with intact chromosome 5q present in del(5q)MDS patients have significant alterations in gene expression despite normal levels of RPS14 (Figure 7D; supplemental Table 6). The commonly upregulated pathways (ROS response and cell cycle regulation pathways) suggest both 5q^+^ and 5q^−^ cells are both responding to oxidative damage. This would indicate that the 5q^−^ cells damage the marrow microenvironment and indirectly impair the effective differentiation of neighboring genetically-normal 5q^+^ cells blocking their optimal expansion. The 5q^+^ cells partially compensate but remain impacted and cannot fully expand and differentiate (Figure 7D,F).

Our studies also provide insight into the posttranscriptional mechanisms that regulate erythropoiesis, such as the alternative splicing of mRNA.^45-47^ In addition to mature spliced transcripts, there was stage-specific and gene-specific intron retention (Figure 3B-C) leading to transcripts unlikely to translate into proteins. There was such a prevalence of intron retention that RNA velocity analysis predicted a reversed trajectory of differentiation after the CFU-E stage (Figure 3A). Besides providing a vehicle to order cells and align studies, the data imply that alternative splicing and intron retention may be especially important regulatory processes in red cell differentiation, as proposed by others.^39,40,45-47^

## Data Availability

Raw data are available via the Dryad data repository at https://doi.org/10.5061/dryad.573n5tb77

## Acknowledgements

We would like to thank our patients, especially patient D1 for their support of this project. This research was funded by NIH grants HL031823 and CA190122 to JLA and QT, respectively, and from Institute for Systems Biology’s Innovator Award Program to CGL. The authors thank Chenkai Luo for early work developing and testing de-multiplexing methods, and Kalyanashis Chakraborty for preliminary analysis of external data sets and integrating them into our analysis pipeline. Human bone marrow samples were obtained at the University of Washington clinical research center which is supported by the NIH National Center for Advancing Translational Sciences, Award Number UL1TR000423.

## Authorship Contributions

RTD, JL, and QT designed the studies; RTD, ADM, ZY, and CGL performed the experiments and generated figures; CGL processed the raw data and developed novel analytical methods; CGL, CM, and XY performed the bioinformatics analysis; RTD and JLA coordinated and directed the studies; RTD, CGL, and JLA wrote the manuscript and all authors edited the manuscript.

## References

1. Santini V. Anemia as the Main Manifestation of Myelodysplastic Syndromes. Semin Hematol. 2015;52(4):348–356.

2. Quigley JG, Gazda H, Yang Z, Ball S, Sieff CA, Abkowitz JL. Investigation of a putative role for FLVCR, a cytoplasmic heme exporter, in Diamond-Blackfan anemia. Blood Cells Mol Dis. 2005;35(2):189–192.

3. Yang Z, Keel SB, Shimamura A, et al. Delayed globin synthesis leads to excess heme and the macrocytic anemia of Diamond Blackfan anemia and del(5q) myelodysplastic syndrome. Sci Transl Med. 2016;8(338):338ra367.

4. Sieff CA, Yang J, Merida-Long LB, Lodish HF. Pathogenesis of the erythroid failure in Diamond Blackfan anaemia. Br J Haematol. 2010;148(4):611–622.

5. Ellis SR. Nucleolar stress in Diamond Blackfan anemia pathophysiology. Biochim Biophys Acta. 2014;1842(6):765–768.

6. Horos R, von Lindern M. Molecular mechanisms of pathology and treatment in Diamond Blackfan Anaemia. Br J Haematol. 2012;159(5):514–527.

7. Narla A, Payne EM, Abayasekara N, et al. L-Leucine improves the anaemia in models of Diamond Blackfan anaemia and the 5q-syndrome in a TP53-independent way. Br J Haematol. 2014;167(4):524–528.

8. Gastou M, Rio S, Dussiot M, et al. The severe phenotype of Diamond-Blackfan anemia is modulated by heat shock protein 70. Blood Advances. 2017;1(22):1959–1976.

9. Rio S, Gastou M, Karboul N, et al. Regulation of globin-heme balance in Diamond-Blackfan anemia by HSP70/GATA1. Blood. 2019;133(12):1358–1370.

10. Gardenghi S, Marongiu MF, Ramos P, et al. Ineffective erythropoiesis in beta-thalassemia is characterized by increased iron absorption mediated by down-regulation of hepcidin and up-regulation of ferroportin. Blood. 2007;109(11):5027–5035.

11. De Franceschi L, Bertoldi M, Matte A, et al. Oxidative stress and beta-thalassemic erythroid cells behind the molecular defect. Oxid Med Cell Longev. 2013;2013:985210.

12. Iskander D, Wang G, Heuston EF, et al. Single-cell profiling of human bone marrow progenitors reveals mechanisms of failing erythropoiesis in Diamond-Blackfan anemia. Sci Transl Med. 2021;13(610):eabf0113.

13. Doty RT, Phelps SR, Shadle C, Sanchez-Bonilla M, Keel SB, Abkowitz JL. Coordinate expression of heme and globin is essential for effective erythropoiesis. J Clin Invest. 2015;125(12):4681–4691.

14. Dixon SJ, Lemberg KM, Lamprecht MR, et al. Ferroptosis: an iron-dependent form of nonapoptotic cell death. Cell. 2012;149(5):1060–1072.

15. Jiang L, Kon N, Li T, et al. Ferroptosis as a p53-mediated activity during tumour suppression. Nature. 2015;520(7545):57–62.

16. Chen JJ. Regulation of protein synthesis by the heme-regulated eIF2alpha kinase: relevance to anemias. Blood. 2007;109(7):2693–2699.

17. Zhang S, Macias-Garcia A, Velazquez J, Paltrinieri E, Kaufman RJ, Chen JJ. HRI coordinates translation by eIF2alphaP and mTORC1 to mitigate ineffective erythropoiesis in mice during iron deficiency. Blood. 2018;131(4):450–461.

18. Zhang J, Socolovsky M, Gross AW, Lodish HF. Role of Ras signaling in erythroid differentiation of mouse fetal liver cells: functional analysis by a flow cytometry-based novel culture system. Blood. 2003;102(12):3938–3946.

19. Hu J, Liu J, Xue F, et al. Isolation and functional characterization of human erythroblasts at distinct stages: implications for understanding of normal and disordered erythropoiesis in vivo. Blood. 2013;121(16):3246–3253.

20. Li J, Hale J, Bhagia P, et al. Isolation and transcriptome analyses of human erythroid progenitors: BFU-E and CFU-E. Blood. 2014;124(24):3636–3645.

21. Brown G, Ceredig R. Modeling the Hematopoietic Landscape. Front Cell Dev Biol. 2019;7:104.

22. Cheng H, Zheng Z, Cheng T. New paradigms on hematopoietic stem cell differentiation. Protein Cell. 2020;11(1):34–44.

23. Zhang Y, Gao S, Xia J, Liu F. Hematopoietic Hierarchy - An Updated Roadmap. Trends Cell Biol. 2018;28(12):976–986.

24. Pellin D, Loperfido M, Baricordi C, et al. A comprehensive single cell transcriptional landscape of human hematopoietic progenitors. Nat Commun. 2019;10(1):2395.

25. Huang P, Zhao Y, Zhong J, et al. Putative regulators for the continuum of erythroid differentiation revealed by single-cell transcriptome of human BM and UCB cells. Proc Natl Acad Sci U S A. 2020;117(23):12868–12876.

26. McInnes L, Healy J, Melville J. UMAP: Uniform Manifold Approximation and Projection for Dimension Reduction. arXiv. 2020;1802.03426.

27. Abkowitz JL, Schaison G, Boulad F, et al. Response of Diamond-Blackfan anemia to metoclopramide: evidence for a role for prolactin in erythropoiesis. Blood. 2002;100(8):2687–2691.

28. La Manno G, Soldatov R, Zeisel A, et al. RNA velocity of single cells. Nature. 2018;560(7719):494–498.

29. Wolf FA, Angerer P, Theis FJ. SCANPY: large-scale single-cell gene expression data analysis. Genome Biol. 2018;19(1):15.

30. Bergen V, Lange M, Peidli S, Wolf FA, Theis FJ. Generalizing RNA velocity to transient cell states through dynamical modeling. Nat Biotechnol. 2020.

31. Xu J, Falconer C, Nguyen Q, et al. Genotype-free demultiplexing of pooled single-cell RNA-seq. Genome Biol. 2019;20(1):290.

32. Huang Y, McCarthy DJ, Stegle O. Vireo: Bayesian demultiplexing of pooled single-cell RNA-seq data without genotype reference. Genome Biol. 2019;20(1):273.

33. Satija R, Farrell JA, Gennert D, Schier AF, Regev A. Spatial reconstruction of single-cell gene expression data. Nat Biotechnol. 2015;33(5):495–502.

34. Traag VA, Waltman L, van Eck NJ. From Louvain to Leiden: guaranteeing well-connected communities. Sci Rep. 2019;9(1):5233.

35. Polanski K, Young MD, Miao Z, Meyer KB, Teichmann SA, Park JE. BBKNN: fast batch alignment of single cell transcriptomes. Bioinformatics. 2020;36(3):964–965.

36. Kuleshov MV, Jones MR, Rouillard AD, et al. Enrichr: a comprehensive gene set enrichment analysis web server 2016 update. Nucleic Acids Res. 2016;44(W1):W90–97.

37. Kidoguchi K, Ogawa M, Karam JD, Martin AG. Augmentation of fetal hemoglobin (HbF) synthesis in culture by human erythropoietic precursors in the marrow and peripheral blood: studies in sickle cell anemia and nonhemoglobinopathic adults. Blood. 1978;52(6):1115–1124.

38. Giarratana MC, Kobari L, Lapillonne H, et al. Ex vivo generation of fully mature human red blood cells from hematopoietic stem cells. Nat Biotechnol. 2005;23(1):69–74.

39. Pimentel H, Parra M, Gee SL, Mohandas N, Pachter L, Conboy JG. A dynamic intron retention program enriched in RNA processing genes regulates gene expression during terminal erythropoiesis. Nucleic Acids Res. 2016;44(2):838–851.

40. Edwards CR, Ritchie W, Wong JJ, et al. A dynamic intron retention program in the mammalian megakaryocyte and erythrocyte lineages. Blood. 2016;127(17):e24–e34.

41. An X, Chen L. Flow Cytometry (FCM) Analysis and Fluorescence-Activated Cell Sorting (FACS) of Erythroid Cells. Methods Mol Biol. 2018;1698:153–174.

42. An X, Schulz VP, Li J, et al. Global transcriptome analyses of human and murine terminal erythroid differentiation. Blood. 2014;123(22):3466–3477.

43. Tusi BK, Wolock SL, Weinreb C, et al. Population snapshots predict early haematopoietic and erythroid hierarchies. Nature. 2018;555(7694):54–60.

44. Lu YC, Sanada C, Xavier-Ferrucio J, et al. The Molecular Signature of Megakaryocyte-Erythroid Progenitors Reveals a Role for the Cell Cycle in Fate Specification. Cell Rep. 2018;25(8):2083–2093 e2084.

45. Pimentel H, Parra M, Gee S, et al. A dynamic alternative splicing program regulates gene expression during terminal erythropoiesis. Nucleic Acids Res. 2014;42(6):4031–4042.

46. Shi L, Lin YH, Sierant MC, et al. Developmental transcriptome analysis of human erythropoiesis. Hum Mol Genet. 2014;23(17):4528–4542.

47. Conboy JG. RNA splicing during terminal erythropoiesis. Curr Opin Hematol. 2017;24(3):215–221.

48. Glader BE, Backer K, Diamond LK. Elevated erythrocyte adenosine deaminase activity in congenital hypoplastic anemia. N Engl J Med. 1983;309(24):1486–1490.

49. Fargo JH, Kratz CP, Giri N, et al. Erythrocyte adenosine deaminase: diagnostic value for Diamond-Blackfan anaemia. Br J Haematol. 2013;160(4):547–554.

50. Liao R, Zheng Y, Liu X, et al. Discovering How Heme Controls Genome Function Through Heme-omics. Cell Rep. 2020;31(13):107832.

51. Doty RT, Yan X, Lausted C, et al. Single-cell analyses demonstrate that a heme–GATA1 feedback loop regulates red cell differentiation. Blood. 2019;133(5):457–469.

52. Doty RT, Yan X, Meng C, Lausted C, Tian Q, Abkowitz JL. Single-cell analysis of erythropoiesis in Rpl11 haploinsufficient mice reveals insight into the pathogenesis of Diamond Blackfan anemia. Exp Hematol. 2021;97:66-78.e66.

53. Danilova N, Gazda HT. Ribosomopathies: how a common root can cause a tree of pathologies. Dis Model Mech. 2015;8(9):1013–1026.

54. Dutt S, Narla A, Lin K, et al. Haploinsufficiency for ribosomal protein genes causes selective activation of p53 in human erythroid progenitor cells. Blood. 2011;117(9):2567–2576.

55. Caceres G, McGraw K, Yip BH, et al. TP53 suppression promotes erythropoiesis in del(5q) MDS, suggesting a targeted therapeutic strategy in lenalidomide-resistant patients. Proc Natl Acad Sci U S A. 2013;110(40):16127–16132.

56. Patel AP, Tirosh I, Trombetta JJ, et al. Single-cell RNA-seq highlights intratumoral heterogeneity in primary glioblastoma. Science. 2014;344(6190):1396–1401.

57. Fiorito V, Allocco AL, Petrillo S, et al. The heme synthesis-export system regulates the tricarboxylic acid cycle flux and oxidative phosphorylation. Cell Rep. 2021;35(11):109252.

58. Quigley JG, Yang Z, Worthington MT, et al. Identification of a human heme exporter that is essential for erythropoiesis. Cell. 2004;118(6):757–766.

59. Keel SB, Doty RT, Yang Z, et al. A heme export protein is required for red blood cell differentiation and iron homeostasis. Science. 2008;319(5864):825–828.

60. Ebert BL, Pretz J, Bosco J, et al. Identification of RPS14 as a 5q-syndrome gene by RNA interference screen. Nature. 2008;451(7176):335–339.

61. McGowan KA, Pang WW, Bhardwaj R, et al. Reduced ribosomal protein gene dosage and p53 activation in low risk myelodysplastic syndrome. Blood. 2011.

62. Narla A, Vlachos A, Nathan DG. Diamond Blackfan anemia treatment: past, present, and future. Semin Hematol. 2011;48(2):117–123.

63. Welch JJ, Watts JA, Vakoc CR, et al. Global regulation of erythroid gene expression by transcription factor GATA-1. Blood. 2004;104(10):3136–3147.

64. Yu M, Riva L, Xie H, et al. Insights into GATA-1-mediated gene activation versus repression via genome-wide chromatin occupancy analysis. Mol Cell. 2009;36(4):682–695.

65. Pfaffl MW. A new mathematical model for relative quantification in real-time RT-PCR. Nucleic Acids Res. 2001;29(9):e45.

66. Wolock SL, Lopez R, Klein AM. Scrublet: Computational Identification of Cell Doublets in Single-Cell Transcriptomic Data. Cell Syst. 2019;8(4):281–291 e289.

67. Heaton H, Talman AM, Knights A, et al. Souporcell: robust clustering of single-cell RNA-seq data by genotype without reference genotypes. Nat Methods. 2020;17(6):615–620.

68. Trapnell C, Cacchiarelli D, Grimsby J, et al. The dynamics and regulators of cell fate decisions are revealed by pseudotemporal ordering of single cells. Nat Biotechnol. 2014;32(4):381–386.

69. Qiu X, Mao Q, Tang Y, et al. Reversed graph embedding resolves complex single-cell trajectories. Nat Methods. 2017;14(10):979–982.

70. Virtanen P, Gommers R, Oliphant TE, et al. SciPy 1.0: fundamental algorithms for scientific computing in Python. Nat Methods. 2020;17(3):261–272.

