## Supplemental Materials for "The transcriptomic landscape of normal and ineffective erythropoiesis at single cell resolution"

**Supplementary Materials**

**Materials and methods**

*Sample acquisition and preparation.* Bone marrow samples were collected after written informed consent according to a University of Washington institutional review board approved protocol. Bone marrow mononuclear cells were cryo-preserved prior to use in this study. The cryo-preserved bone marrow mononuclear cells were thawed, resuspended in equivalent volume of FBS (Sigma), washed in HBSS, then resuspended at $10^7$ cells per ml in PBS, 0.1% BSA, 2 mM EDTA and incubated with 1:10 dilution of biotinylated CD3 (555338, BD Biosciences), CD19 (555411), and CD11b (555387). Antibody-bound lineage positive cells were depleted with Dynabeads Biotin Binder (Invitrogen) according to the manufacturer's protocol. This rapid column-based depletion of CD3, CD11b, and CD19 expressing bone marrow mononuclear cells resulted in an average 68% increase in lineage negative cells. This minimized cell processing time and significantly enriched the erythroid cell population, while retaining enough non-erythroid cells to enable accurate lineage clustering. As there were insufficient numbers of early erythroid precursors for robust analysis present in uncultured marrow, we also expanded erythroid precursors in culture for 3-6 days. Lineage-depleted marrow mononuclear cells were cultured in step 1 erythroid expansion media as before:[3] IMDM, 5% pooled Human AB plasma (Innovative Research), 330 μg/ml holo-transferrin, 1 μM dexamethasone, 160 mM monothioglycerol (Sigma), 10 μg/ml Insulin (Humulin R, Lily), 2 U/ml preservative free heparin (Pfizer labs), 2 U/ml EPO (Procrit, Janssen), 100 ng/ml SCF and 5 ng/ml IL3, (Peprotech) at 2 x $10^5$ cells/ml. The cells were re-seeded at 2 x $10^5$ cells/ml on day 3 with fresh media.

*Flow cytometry.* Approximately $10^4$ cells were stained with 5 μg/ml of CD3-PE/Cy7, CD19-PE/Cy7, CD11b-PE/Cy7, CD36-Pacific Blue, CD71-APC (transferrin receptor), CD235a-PE (Glycophorin A) (all from BD biosciences) for 30 minutes. Washed cells were resuspended in PBS with 1% BSA and 0.1% sodium azide at 2.5 x $10^5$/ml. Flow cytometry data was collected on a LSR II cytometer and data was analyzed with FlowJo (v10, BD Biosciences).

*Quantitative PCR*. Ex vivo marrow (day 0) which was predominantly late erythroid cells, and cultured cells (day 6) which were predominantly early stage erythroid cells, were lysed in Trizol and mRNA purified according to manufacturer's instructions. First strand cDNA synthesis was performed with iScript (Bio-

Rad) and analyzed by quantitative PCR using *ACTB* as the reference gene[65]. Primers and probes are listed in supplemental Table 7.

*Antibody barcoding.* Cells were washed and resuspended in PBS with 1% BSA at 5 x $10^5$/ml and incubated with 10 μg/ml of each barcoded antibody: CD3, CD11b, CD14, CD36, CD70, CD71/TfR, CD117/cKit, CD235a/GlyA. SPARTA antibodies were from BD Biosciences and barcoded in house using the SoluLinK benzaldehyde conjugation kit (Trilink Biotechnologies #S-9011-1) according to the manufacturer's instructions. CITE-seq barcoded antibodies were obtained from BioLegend. Once cells were labeled by antibody-mediated barcoding, further purification steps were not needed, reducing processing time and minimizing cell manipulations that could damage the quality of the mRNA and cDNA libraries.

*Sample processing.* We generated 41 scRNAseq samples from 7 normal individuals, 2 DBA patients, and 2 del(5q) MDS patients. Single-cell mRNA barcoding and library preparation were performed on the 10x Chromium controller according to the manufacturer's instructions. A single GEM (gel beads-in-emulsion) well from each sample was used to generate both the gene expression and antibody capture libraries which were recombined for sequencing. The libraries were sequenced on a NextSeq 550 or Novaseq 6000 instrument and raw sequence data was processed with Cell Ranger. We sequenced 74,214 individual cells. Doublets were filtered out using Scrublet (v.0.2.2).[66] Cells containing more than 15% mitochondrial reads, which indicates high plasma membrane permeability, were filtered out (4869 removed, 69345 retained) as dead. We chose a 15% mitochondrial read cutoff to maximize inclusion of terminally differentiating erythroid precursors while still filtering out dead cells. Genes not appearing in at least 20 cells were filtered out (14711 removed, 18827 retained). Second, a list of the 7500 most highly-variable genes was determined, taking the variation of the five experiments into consideration (*pp.highly_variable_genes* function, setting the "batch_key" to "experiment"). To this list, additional key progenitor, erythroid-lineage, and proliferation genes were added (*HBB, HBA1, GYPA, GYPB, GATA1, TAL1, KLF1, KLF3, ZFPM1, EIF2AK1, E2F2, TFRC, SLC25A37, SLC48A1, ALAS2, HMBS, FECH, TOP2A, KIF11, HEMGN, RPL35, RPL8, RPL4, RPL5, RPL11, RPS14, RPS19, MRPL57, MRPL40, CDKN1A, SLC7A11, GPX4, PRDX2, GPX1, COX6B1, CYC1, NDUFA8, NDUFB6, SDHB, PSMA2, PSMB2, SMC2, KIF22, KIF2A, SMC1A, SMC4, KIF2C, AURKA, AURKB, GCLM, FTH1, FTL, MAF1,*

*KEAP1, SLC52A2, SLC20A1, SLC20A2, XPR1, FLVCR1, SLC5A6, SLC1A5, SLC2A1, SLC4A1, KIT, CD36, TRIB2, CA1, CA2, SPINK2*). A total of 7545 genes were retained.

*Data aggregation*. FASTQ file reads were aligned to the human genome (GRCh38) by CellRanger (v.3.0) and the results saved to bam files which were analyzed by Velocyto and transcript splice quantification saved as loom files. Loom files were aggregated by ScanPy[29] (*AnnData.concatenate* function). Transcript counts were normalized to $1\times10^4$ UMIs per cell and log-transformed (*pp.log1p* function). Each gene was scaled to unit variance (*pp.scale* function). The aggregated data was saved to a single HDF5 file ("step01_aggr_5exp_211012.h5ad").

*Sample demultiplexing.* Samples in Experiments 1 and 2 were analyzed with separate 10x Genomics bead libraries and required no demultiplexing. Samples in Experiments 3-5 were performed multiplex, with each bead library containing both an anemia and normal control samples. Samples were demultiplexed based on natural genetic variation and without the assistance of donor genome references using Vireo,[32] ScSplit,[31] and Souporcell,[67] with the three methods providing nearly identical results using default settings. Vireo (v0.2.2) was selected based on its speed and its file formats. Multiplexed sample identity was confirmed by checking for the presence of the RPS19-L18P mutation (Experiment 3), for attenuated RPS14 gene expression due to the chromosome 5q deletion (Experiments 4-5), and sex-specific (*XIST, RPS4Y1*) gene expression (Experiment 5).

*Del(5q) MDS sample deconvolution.* We took two approaches to determine which cells in the MDS patient had the 5q deletion and which had intact chromosome 5q. First, we used gene expression data across all chromosomes to identify cells with the 5q13-33 deletion according to the method of Patel et al.[56] This approached identified 75% of marrow cells from patient M1 as containing the 5q13-33 deletion which closely match the iFISH results. Second, we took advantage of coordinate expression levels of ribosomal proteins and compared the expression level of *RPS14* (within the deletion interval) to the expression level of *RPS18* (Ch6p21.32) which is expressed at comparable levels in normal cells. Using an RPS14:RPS18 ratio threshold of 0.75, 77% of erythroid cells were identified as expressing haploinsufficient levels of RPS14. The two methods produced good agreement with 91.1% concordance of the cell assignments (supplemental Figure 7). As ribosomal protein genes are downregulated along with all other non-erythroid-specific genes during terminal erythropoiesis, resolution of $5q^+$ vs $5q^-$ cells via ribosomal protein gene expression past the basophilic erythroblast stage is unreliable.

*Cell typing and visualization.* Gene set scores were calculated using the ScanPy *score_genes* function. Eleven cell type scores were calculated using panels of six marker genes: erythrocyte (*HBA1, CA1, SLC4A1, GYPA, TFRC, HMGA1*), multipotent progenitor (*KIT, CCL2, GATA2, GATA4, RUNX1, TRIM33*), common lymphoid progenitor (*SPINK2, DNTT, CD79A, EBF1, IGHM*), T cell (*CD3D, TRAC, CD8A, CD4, CXCR4, BTG1*), natural killer (*GNLY, NKG7, PRF1, KLRB1, GZMA, CTSW*), B cell (*CD27, CD74, CD79B, IGKC, IGHM, MZB1*), dendritic cell (*PLD4, ITM2C, JCHAIN, CD40, PLIN2, LGALS3BP*), macrophage (*TYROBP, FCER1G, CD14, S100A8, S100A9, LYZ*), and neutrophil (*PRSS57, SMIM24, ELANE, AZU1, PRTN3, CFD*). Scores were also calculated for S and G2/M cell cycle phase using the gene sets and methods of Satija et al.[33] Principal components were calculated using the 2500 most variable genes (*pp.highly_variable_genes* function, setting the “batch_key” to “phase”). Cells were clustered by Leiden community detection[34] using batch-corrected nearest neighbors.[35] Nearest neighbors were calculated using the top 25 principal components. A batch consisted of an experiment with both normal and anemic marrows (each experiment) and a culture period (zero, three, or six days). Clusters were labeled as specific cell types based on the comparative cell type scores. Cell clusters were visualized using two-dimensional UMAP (Uniform Manifold Approximation and Projection).[26] Based on the UMAP, the erythrocyte clusters were divided into two subtypes. The first subtype, labeled "Ery1", includes the earlier stages of differentiation which were expanded in 6 days of culture. The second subtype, "Ery2", represents later stages of erythroid development comprised of uncultured marrow predominately from healthy individuals and contains cells expressing higher levels of hemoglobin genes and fewer total genes as expected during terminal erythroid differentiation.

We identified 9 distinct cell-type clusters in all samples which were assigned to cell lineages consistent with protein expression (Figure 1B-C, Supplemental Figure 2A-B). Culture increased the numbers of early and intermediate erythroid precursors while decreasing the frequency of other lineages and late stage erythroid precursors that are prevalent in marrow (Figure 1D-E). By day 6, there were sufficient early and intermediate stage precursor cells for comprehensive analysis of both normal and ineffective erythropoiesis as expected.[38] Differentiation progressed from MPP to erythroblasts evident by gene expression of individual cells and consistent with traditional flow cytometric criteria[18-20] (Figure 1D; supplemental Figure 1). Both normal and patient cultured cells had increased expression of globin genes (eg. *HBG1*) and *TFRC* (supplemental Figure 3C) as previously reported.[37,38]

*Pseudotemporal ordering.* Spliced and unspliced transcript moments and transcript velocities were calculated using scVelo[30] and the *pp.moments* and *tl.velocity* functions. As unspliced mRNA precedes mature spliced mRNA, Velocyto and scVelo use changes in the ratio of spliced to unspliced mRNA to predict developmental trajectories and allowed consistent alignment across distinct experiments unlike other trajectory algorithms such as monocle[68,69] which worked well within each experiment but did not allow uniform alignment of combined data from distinct experiments. Pseudotemporal ordering of the erythroid-lineage cells was determined using the *tl.velocity_pseudotime* function separately on Ery1 and Ery2. Velocity pseudotime (VPT) from 0 to 1 was calculated for Ery1 cells using the cell with the highest *KIT* expression as the root (VPT=0.0) and the cell with the highest *HBA1* as the terminus (VPT=1.0). VPT from 1 to 2 was calculated for Ery2 using the cell with the maximum number of expressed genes as the root (VPT=1.0) and the cell with the fewest expressed genes as the terminus (VPT=2.0). Based on VPT and CITEseq, erythropoiesis was divided into seven stages, labeled A through F, with cut points at 0.00, 0.10, 0.25, 0.70, 0.95, 1.40, 1.70, and 2.00, respectively. Stage C was divided into two substages, with C1 from 0.25 - 0.4 and C2 from 0.4 - 0.70 which matched transcriptional changes within the CFU-E population.

*UMAP visualization.* Cell annotations, gene expression levels, cell densities, and cell velocities were visualized on two-dimensional projections calculated from manifold learning (tl.umap function). For cell density plots (pl.embedding_density function), cells were deemed positive for gene or protein if they expressed greater than the median level for all cells. ScVelo was used to project (tl.velocity_embedding function) and display (pl.velocity_embedding_stream function) single-cell velocities on UMAP plots.

*Alternate erythroid trajectories.* Erythroid-lineage cells (Ery1 and Ery2) were visualized by diffusion map (*tl.diffmap* function). Diffusion maps are used to visualize the continuous process of erythroid differentiation in 2 dimensional space, while scatter plots are used to visualize changes in individual genes or pathways over pseudotime. The visualizations suggested two trajectories: a primary continuum of cells ranging from VPT=0 to VPT=2, as well as a second trajectory branching off at approximately

VPT=0.35 and terminating around VPT=0.75. These were labeled trajectory A and trajectory B, respectively. Trajectory B was defined as Leiden cluster 10 from the clustering of all cell types.

*Statistical analysis: Differential expression and gene set enrichment.* Genes, gene sets, and proteins were tested for differential expression between two groups of cells using the Wilcoxon rank-sum test with Benjamini-Hochberg correction. Gene measurements stored in the sparse matrix format were tested using ScanPy (*tl.rank_genes_groups* function, setting “method” to “wilcoxon”; P<0.05 ), while measurements stored in dense matrices were tested using SciPy (stats.mannwhitneyu function, setting “alternative” to “two-sided”; P<0.05).[29,70] GO-term enrichment analysis was performed on the DEG using the Enrichr method[36] as implemented by the GSEApy software library. The gene data set used was downloaded from the Molecular Signatures Database (version 7.4, http://www.gsea-msigdb.org/gsea/msigdb/). Enrichr calculates both the odds ratio (OR) and the Benjamini-Hochberg corrected P-value. The OR = [Given list in/out] / [expected in/out]. Top-scoring gene sets were plotted as scatter plots, ranked by OR, with bubble sizes proportional to the number of DEG appearing in the gene set and the bubble color representing the corrected P-value.

*Projection of external data.* Single-cell transcriptomic data from the nine samples of Iskander et al.[12] totaling 41,415 cells were imported and normalized as described above. Transcript counts were normalized to $1\times10^4$ UMIs per cell and log-transformed (pp.log1p function), and each gene was scaled to unit variance. Cell lineage marker panels were calculated as before (tl.score_genes function). The two data sets were merged and genes not appearing in both data sets were removed, retaining 6568 genes. UMAP and PCA embeddings, as well as lineage, cluster, vpt, stage, and trajectory annotations were mapped onto the imported data using the ScanPy Ingest method (tl.ingest function). Non-erythroid cells were removed based on the "lineage" mapping. Additional non-erythroid cells were removed that exhibited high lymphoid progenitor scores (CLP > -0.4). For the external-data erythroid cells, a new PCA (pp.pca function), K-nearest neighbor set (bbknn.bbknn function), and diffusion map (pl.diffmap) was calculated. Two branches were determined for the diffusion map using diffusion pseudotime branching (tl.dpt function setting "branches" to "1" and "n_dcs" to "3"). This resulted in three groups of cells: an ambiguous root group, a trajectory A-overlapping group, and a trajectory B-overlapping group.

**Supplemental References**

64. Pfaffl MW. A new mathematical model for relative quantification in real-time RT-PCR. *Nucleic Acids Res*. 2001;29(9):e45.

65. Wolock SL, Lopez R, Klein AM. Scrublet: Computational Identification of Cell Doublets in Single-Cell Transcriptomic Data. *Cell Syst*. 2019;8(4):281-291 e289.

66. Heaton H, Talman AM, Knights A, et al. Souporcell: robust clustering of single-cell RNA-seq data by genotype without reference genotypes. *Nat Methods*. 2020;17(6):615-620.

67. Trapnell C, Cacchiarelli D, Grimsby J, et al. The dynamics and regulators of cell fate decisions are revealed by pseudotemporal ordering of single cells. *Nat Biotechnol*. 2014;32(4):381-386.

68. Qiu X, Mao Q, Tang Y, et al. Reversed graph embedding resolves complex single-cell trajectories. *Nat Methods*. 2017;14(10):979-982.

69. Virtanen P, Gommers R, Oliphant TE, et al. SciPy 1.0: fundamental algorithms for scientific computing in Python. *Nat Methods*. 2020;17(3):261-272.

**Supplemental Table files:**

Supplemental Table 1, Ery1 DEG.xlsx

Supplemental Table 2, DEG Trajectory B vs A.xlsx

Supplemental Table 3, DEG DBA vs normal.xlsx

Supplemental Table 4, DEG del5qMDS vs normal.xlsx

Supplemental Table 5, DEG 5q-MDS vs normal.xlsx

Supplemental Table 6, DEG 5q+MDS vs normal.xlsx

**Supplemental Table 7. qPCR probe sets.**

| | | | |
|---|---|---|---|
| *ACTB* | NM_001101 | Primer 1 | ATCACGATGCCAGTGGTA |
| | | Primer 2 | AGATGACCCAGATCATGTTTG |
| | | Probe | ACGTTGCTATCCAGGCTGTGCTA |
| *DDX39B* | NM_004640 | Primer 1 | CCGATGAGAATGATGCCAAGA |
| (Spliced) | | Primer 2 | GTCTGTTCAATGTAGGAGGAGATG |
| | | Probe | ATGTGCAGGATCGCTTTGAGGTCA |
| *DDX39B* | NT_167249 | Primer 1 | CCGATGAGAATGATGCCAAGA |
| (Unspliced) | | Primer 2 | GTTTCATGAGATCAGTACTCAC |
| | | Probe | ATGTGCAGGATCGCTTTGAGGTCA |
| *GATA1* | NM_002049 | Primer 1 | AGATGAATGGGCAGAACAGG |
| (Spliced) | | Primer 2 | ATTTCTCCGCCACAGTGTC |
| | | Probe | TCAGTAAACGGGCAGGTACTCAGTG |
| *GATA1* | NG_008846 | Primer 1 | CCAGGGCACTGATCTCACAT |
| (Unspliced) | | Primer 2 | ATTTCTCCGCCACAGTGTC |
| | | Probe | TCAGTAAACGGGCAGGTACTCAGTG |
| *TFRC* | NM_003234 | Primer 1 | TTTCCACCATCTCGGTCATC |
| (Spliced) | | Primer 2 | GGGACAGTCTCCTTCCATATTC |
| | | Probe | CAGACAATCTCCAGAGCTGCTGCA |
| *TFRC* | NG_046395 | Primer 1 | TTTCCACCATCTCGGTCATC |
| (Unspliced) | | Primer 2 | ATGCAAGACCGCTTTCAAAT |
| | | Probe | CAGACAATCTCCAGAGCTGCTGCA |

### Supplementary Figures

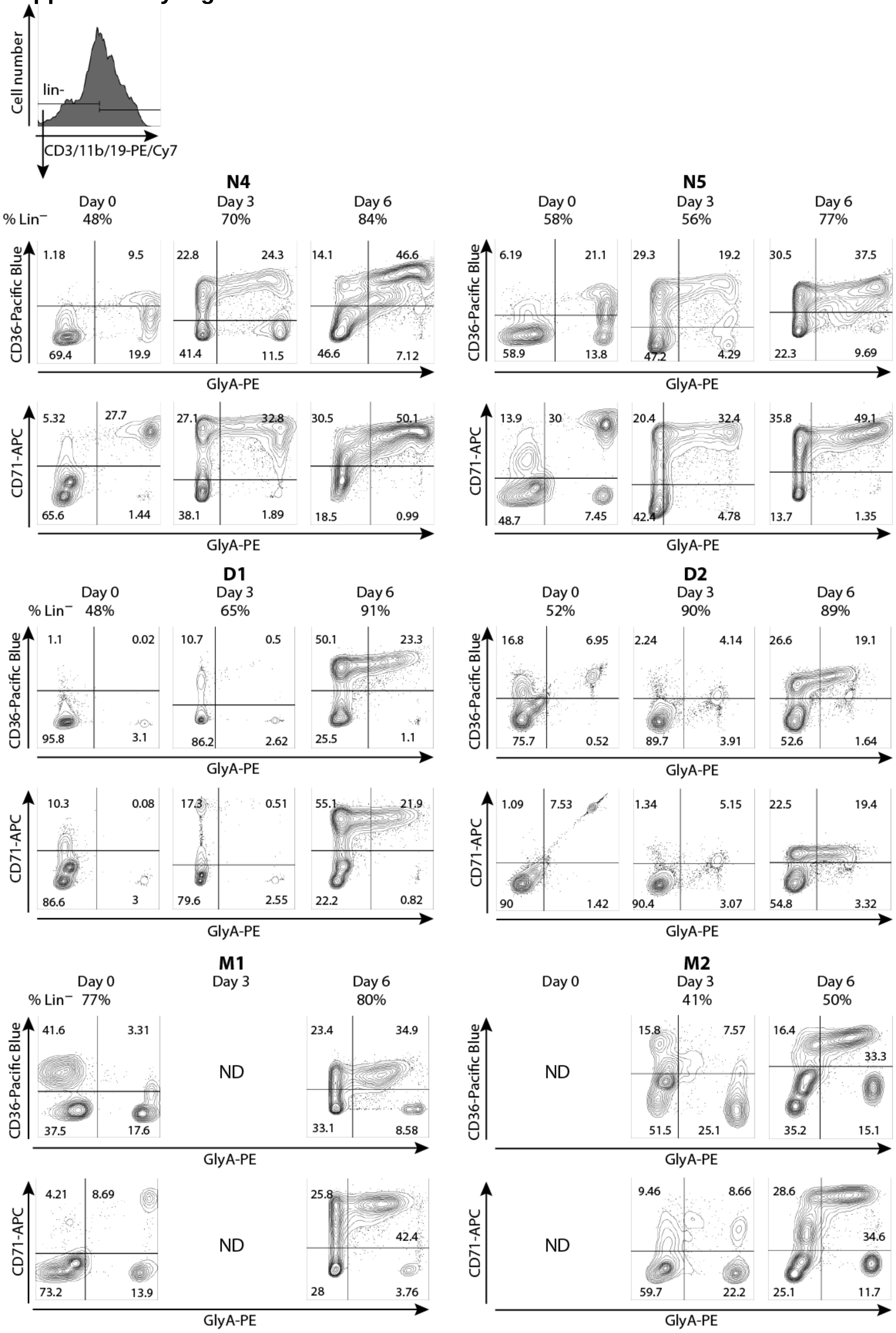


**Supplemental Figure 1. Flow cytometry analysis of lineage negative bone marrow cells.** Bone marrow cells from normal volunteers (N4 & N5), DBA patients (D1 & D2), and the del(5q) MDS patients (M1 & M2) were stained with CD3, CD11b, CD19, CD36, CD71, and CD235a (GlyA). The lineage negative ($CD3^-$ $CD11b^-$ $CD19^-$) cells are presented as CD36 by GlyA and CD71 by GlyA contour plots to show progression of erythroid differentiation during 6 days in culture. The numbers of the most mature cells ($GlyA^+CD36^-$ or $GlyA^+CD71^-$) present in ex vivo marrow decreased during the 6 day culture. ND= not done due to insufficient M1 & M2 cells for flow cytometry analysis.

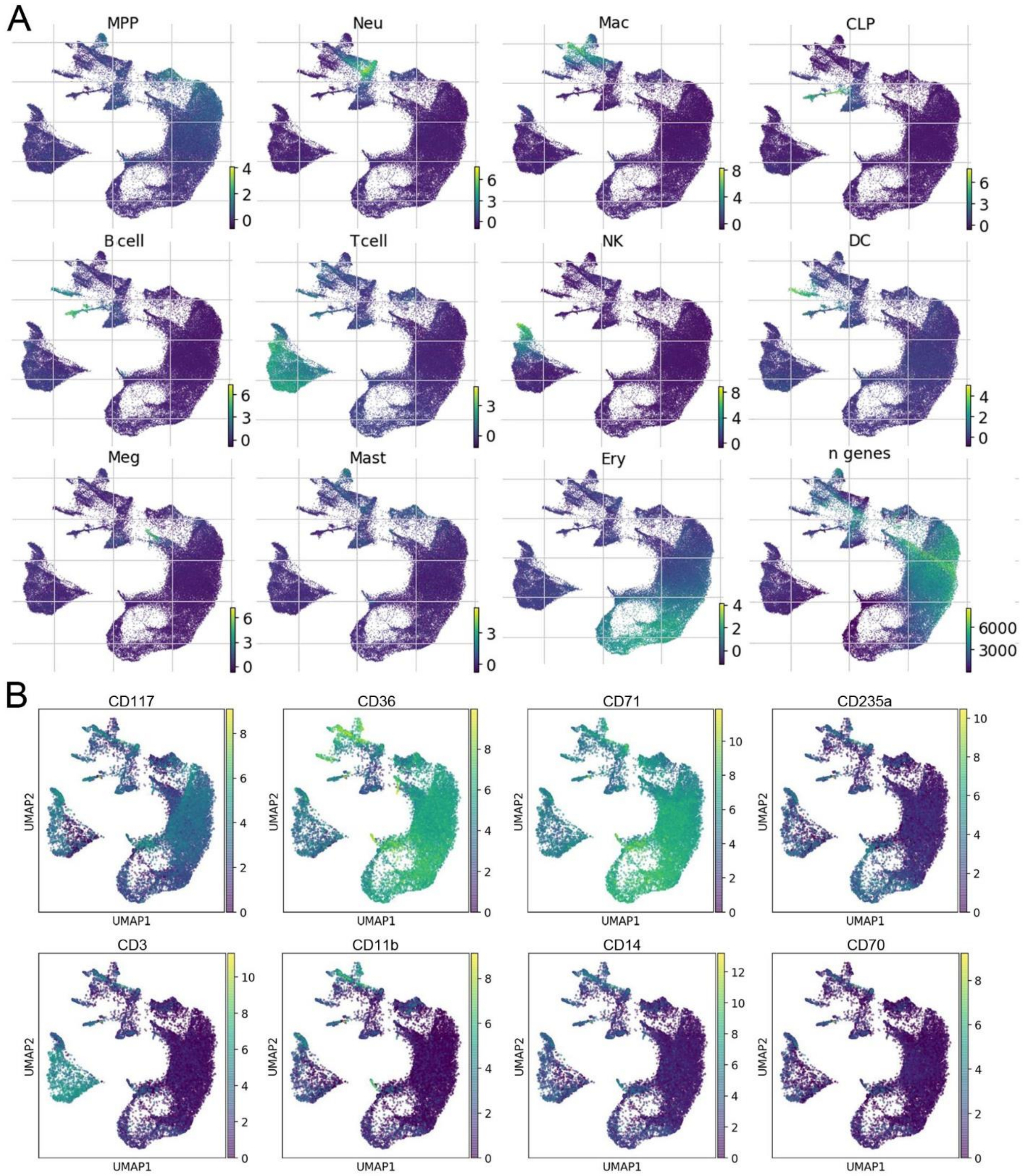


**Supplemental Figure 2. Expression of lineage panel genes and proteins in marrow cells.** (**A**) UMAP visualizations of all bone marrow cells showing the relative expression of globally-distinguishing lineage restricted gene sets (see Figure 1C for genes), and the total number of genes expressed. (**B**) Relative expression of cell surface proteins detected with barcoded antibodies in all marrow cells confirms cell lineages and progression of erythroid differentiation.

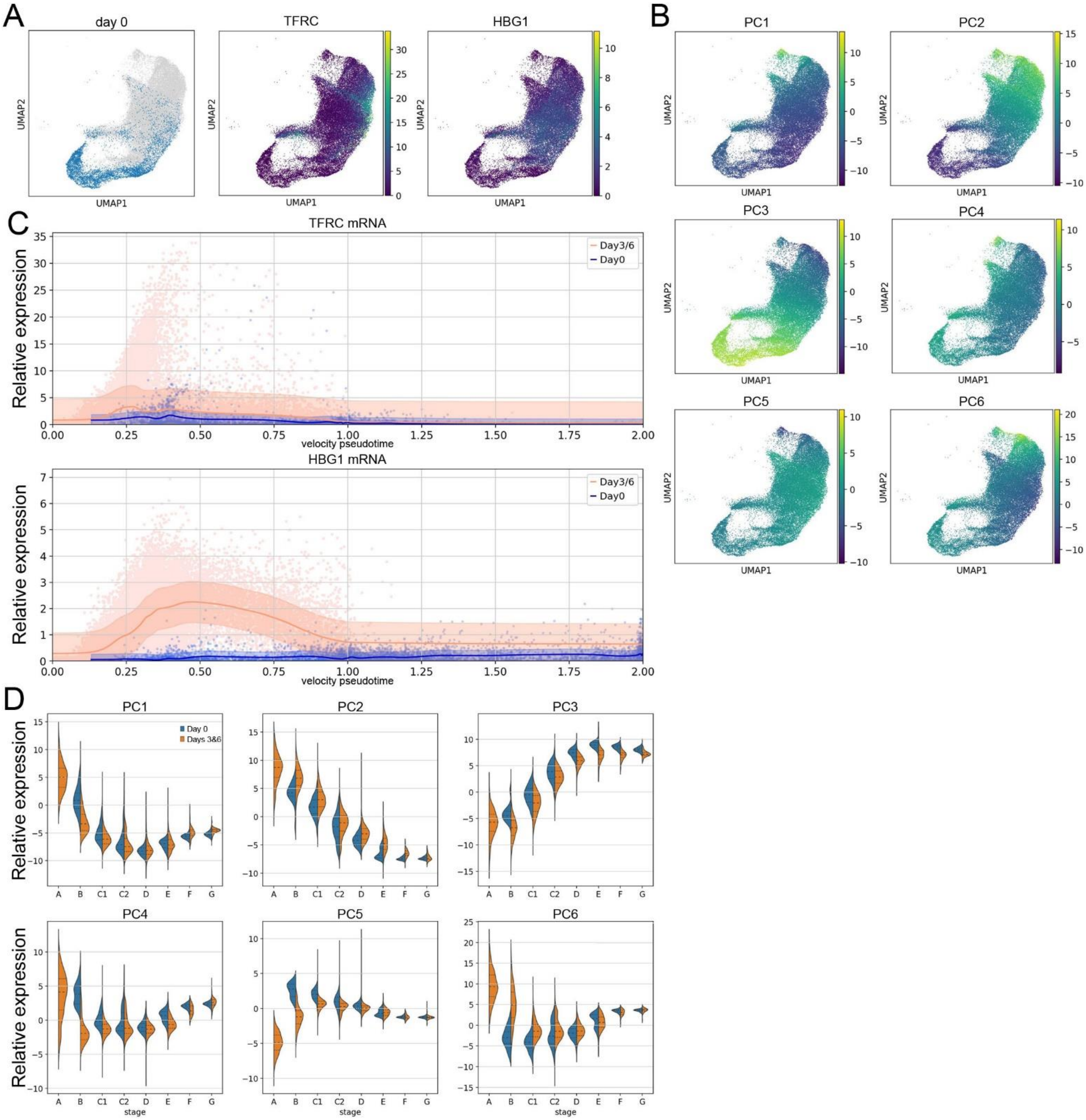


**Supplemental Figure 3. Erythroid gene expression in vitro mirrors erythroid gene expression in vivo.** (**A**) UMAP of erythroid marrow cells highlighting uncultured cells (Day 0) along with relative expression levels of *TFRC* and *HBG1* genes showing upregulation in cultured cells consistent with previous reports.[37,38] (**B**) UMAP of erythroid marrow cells overlaid with relative expression of principal components (PC) 1 to 6 showing subtle differences in cultured cells in the early precursors (top right area in each UMAP). (**C**) Velocity pseudotime plot showing expression of *TFRC* and *HBG1* in uncultured (Day 0) and cultured (Day 3/6) marrow erythroid cells. Mean expression over pseudotime is shown with trend lines with a 1 SD variation highlighted. (**D**) Violin plots of PC 1-6 separated by erythroid stages show minor differences in gene expression during BFU-E and early CFU-E stages. These differences were minor and seem to reflect kinetic differences which resolve by the late CFU-E stage. These minor differences did not interfere with subsequent analysis when cells from day 0, 3, & 6 were combined.

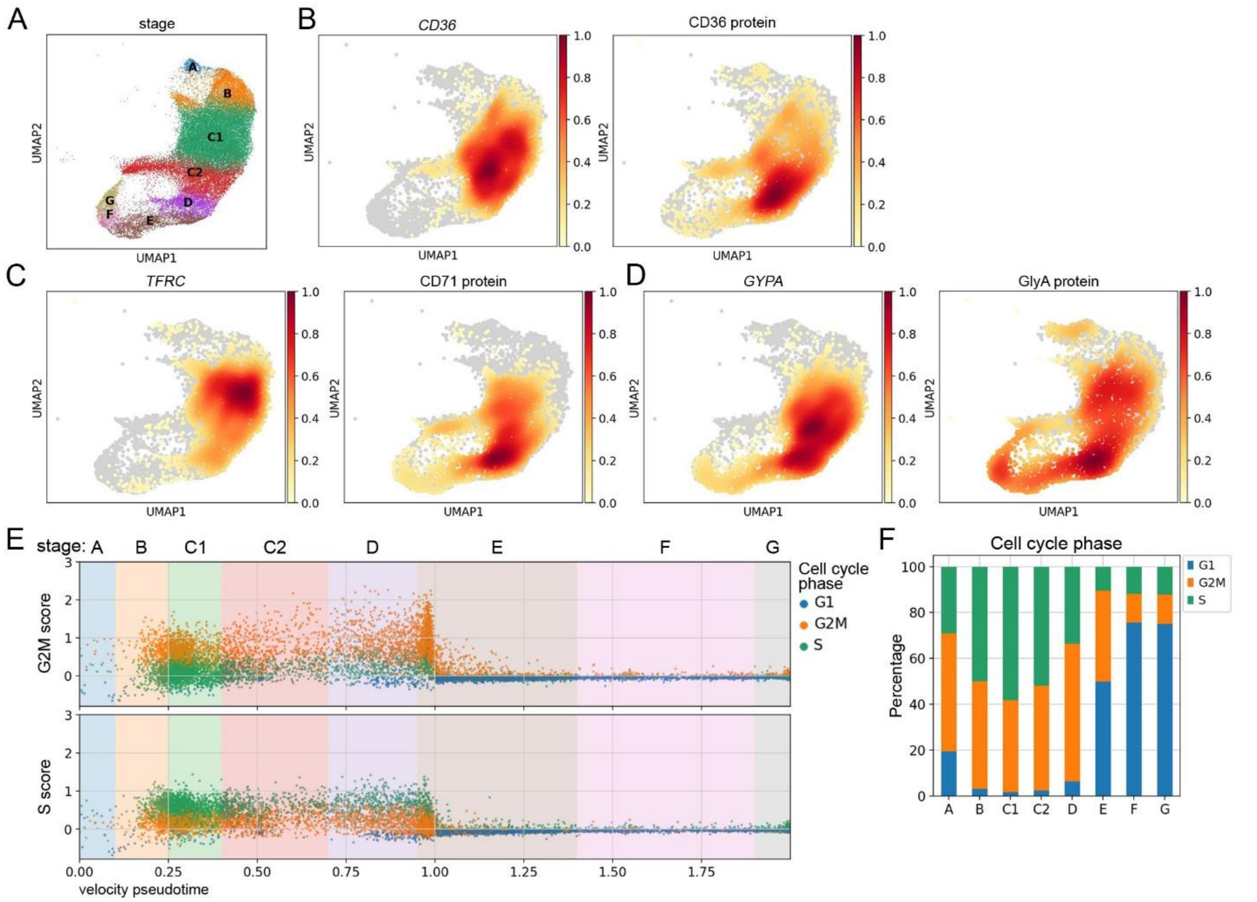


**Supplemental Figure 4. Pseudotime staging of erythroid precursor cells.** (**A**) UMAP visualization of all erythroid progenitor cells with each cell identified by predicted stage of differentiation as described in Figure 2. UMAP visualization of erythroid precursor cells with expression density plots showing the frequency of cells at each position expressing (**B**) CD36 mRNA and protein, (**C**) TFRC/CD71 mRNA and protein and (**D**) GlyA/CD235a mRNA and protein. (**E**) Velocity pseudotime of erythroid progenitors showing the G2M score or S score of individual cells categorized by their assigned cell cycle phase. (**F**) Plots of each stage of differentiation showing the frequency of cells in G1, G2M, or S phase of the cell cycle. Analysis of cell cycle gene expression shows increased rate of cycling at the BFU-E stage with peak frequency of S phase cells at the early CFU-E (C1) stage, consistent with the accelerated cell cycling of CFU-E described by others.[43,44] During the basophilic stage (stage E), there is a dramatic decrease in G2M and S phase gene expression, supporting the concept that early and late basophilic erythroblasts can be distinguished by their distinct surface and transcriptional states.[19,25,41,42]

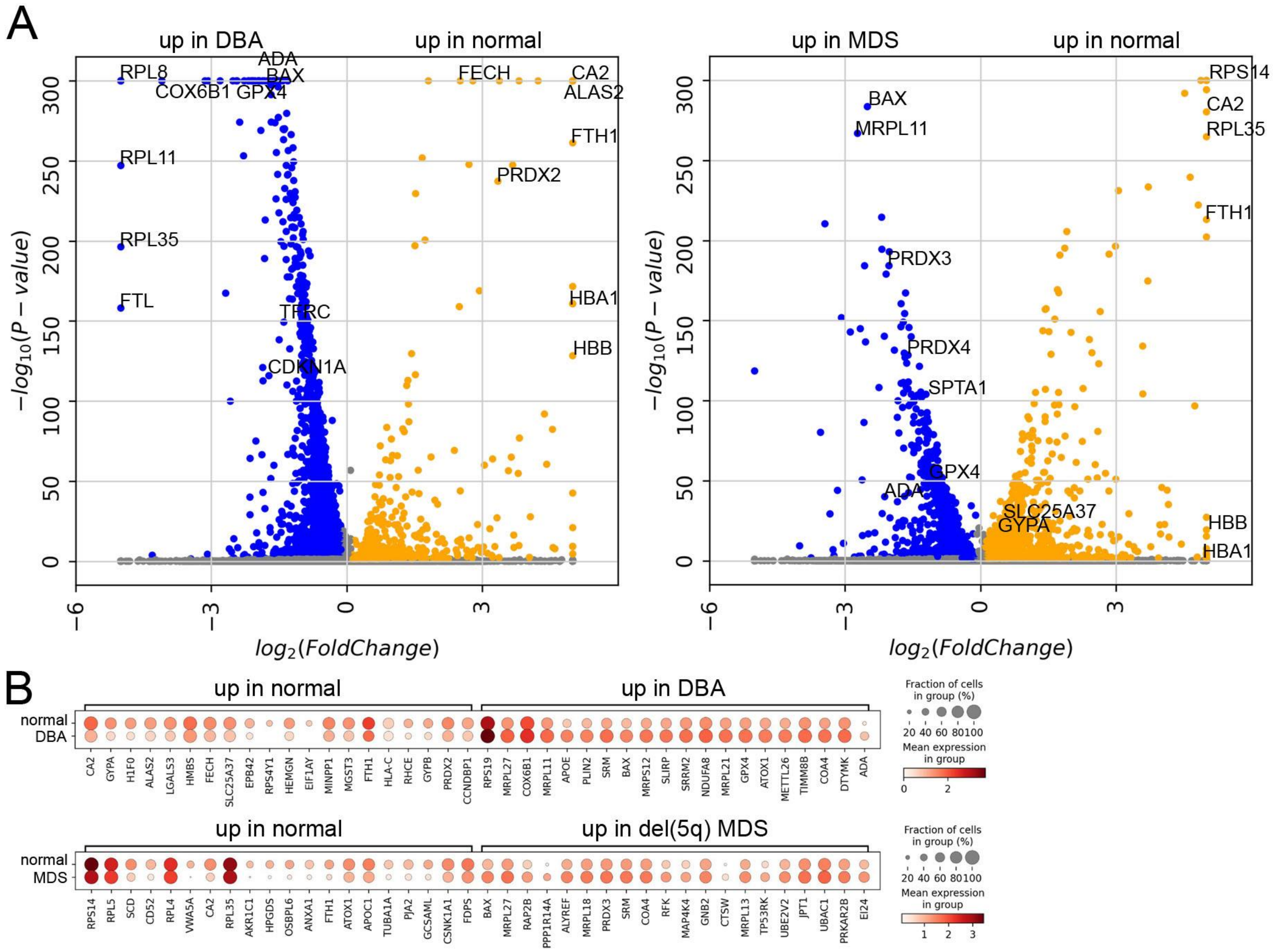


**Supplemental Figure 5. Erythroid precursor cells from DBA and del(5q) MDS anemia patients upregulate stress response, apoptosis, and mitochondrial metabolism genes.** (**A**) Volcano plots showing differentially expressed genes which are upregulated in early erythroid cells (group Ery1, Figure 1) from DBA or del(5q) MDS anemia patients compared to those from normal individuals. Later stage cells (group Ery2, Figure 1) are not present in sufficient numbers from anemia patient samples and are excluded from the analysis. (**B**) Dot plots showing the top 20 genes upregulated in DBA or del(5q) MDS anemia patient marrow erythroid cells (group Ery1) versus those upregulated in erythroid cells from normal individuals. See supplemental Table 1 for complete DEG lists.

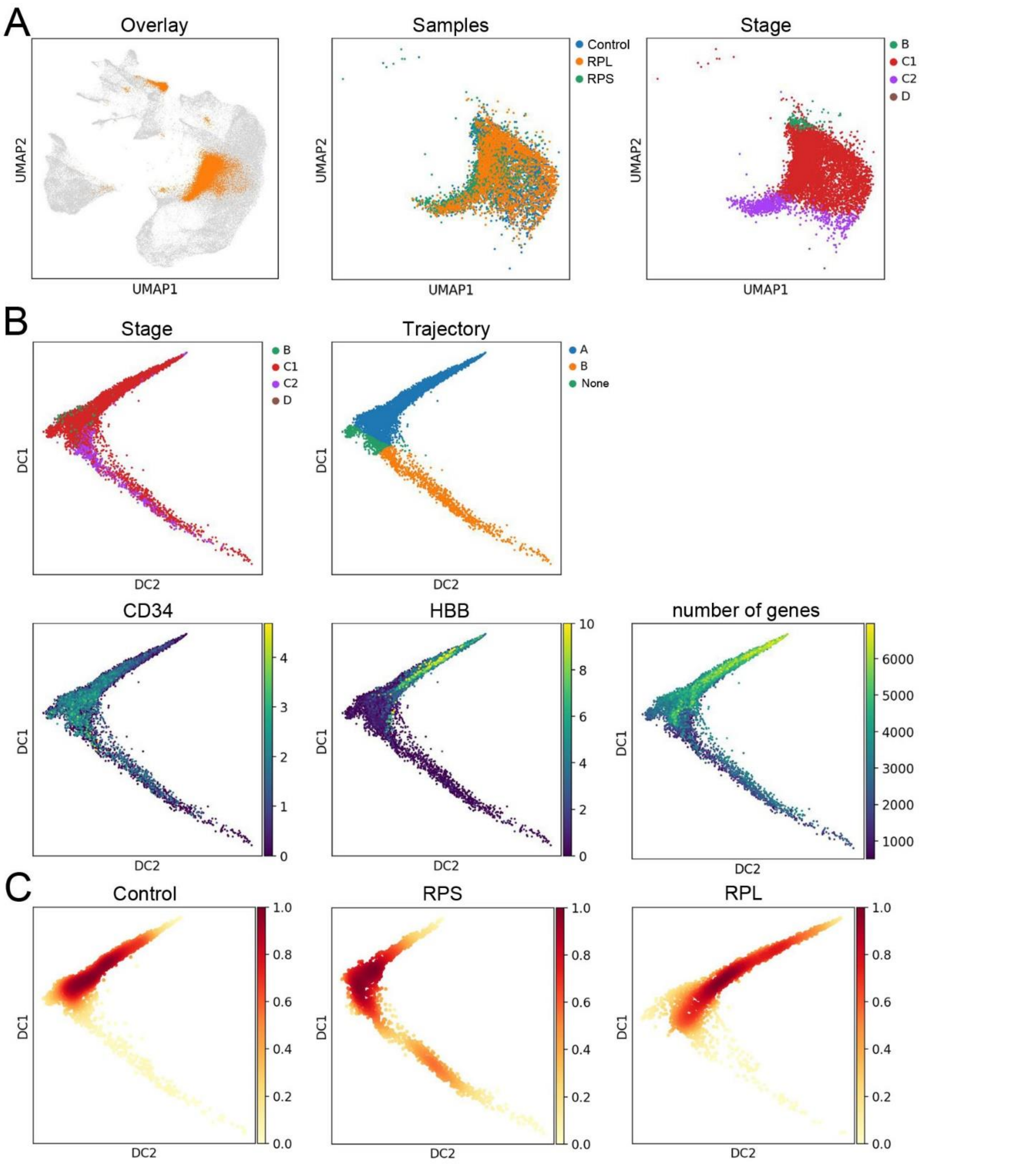


D

| ID | Sample | Trajectory A | Trajectory B | % on B | Mean |
|---|---|---|---|---|---|
| GSM4731561 | control | 1107 | 7 | 0.6% | |
| GSM4731562 | control | 958 | 17 | 1.7% | 8.2±10.0% |
| GSM4731563 | control | 721 | 207 | 22.3% | |
| GSM4731564 | RPS | 240 | 73 | 23.3% | |
| GSM4731565 | RPS | 441 | 259 | 37.0% | 32.1±6.2% |
| GSM4731566 | RPS | 580 | 325 | 35.9% | |
| GSM4731567 | RPL | 436 | 77 | 15.0% | |
| GSM4731568 | RPL | 726 | 58 | 7.4% | 7.8±5.7% |
| GSM4731569 | RPL | 972 | 11 | 1.1% | |

**Supplemental Figure 6. Erythroid precursor cells from DBA and normal individuals follow both trajectories.** (**A**) Cells from Iskander et al[12] overlaid onto our UMAP of all cells (left) using the ingest function while just those cells cells identified as erythroid precursors are presented in the right two panels and then overlaid with the sample type (control, RPL mutations, or RPS mutations) and erythroid stages. The majority of the erythroid cells are identified as CFU-E as expected for marrow CD34$^{+}$ cells. (**B**) Diffusion map visualization of CD34$^{+}$ erythroid precursor cells reveals two distinct trajectories. The cell stage and trajectory assignments are shown. Both trajectories had decreasing *CD34* expression while only trajectory A cells highly upregulated *HBB* and had high levels of gene expression, thus matching trajectories A and B in our samples. (**C**) Diffusion maps showing the density and distribution of individual cells along both trajectories from each sample type. (**D**) The cell counts and percentages of CD34$^{+}$ erythroid precursor cells following each trajectory is consistent with RPS mutations having more severe anemia RPL mutations[12] and suggests that the more cells which follow trajectory B correlates with the severity of anemia. Additionally, cells from healthy individuals follow both trajectories, indicating that erythroid cell precursor cell death on trajectory B is a universal finding. The random patient sample identifiers are from the published study[12].

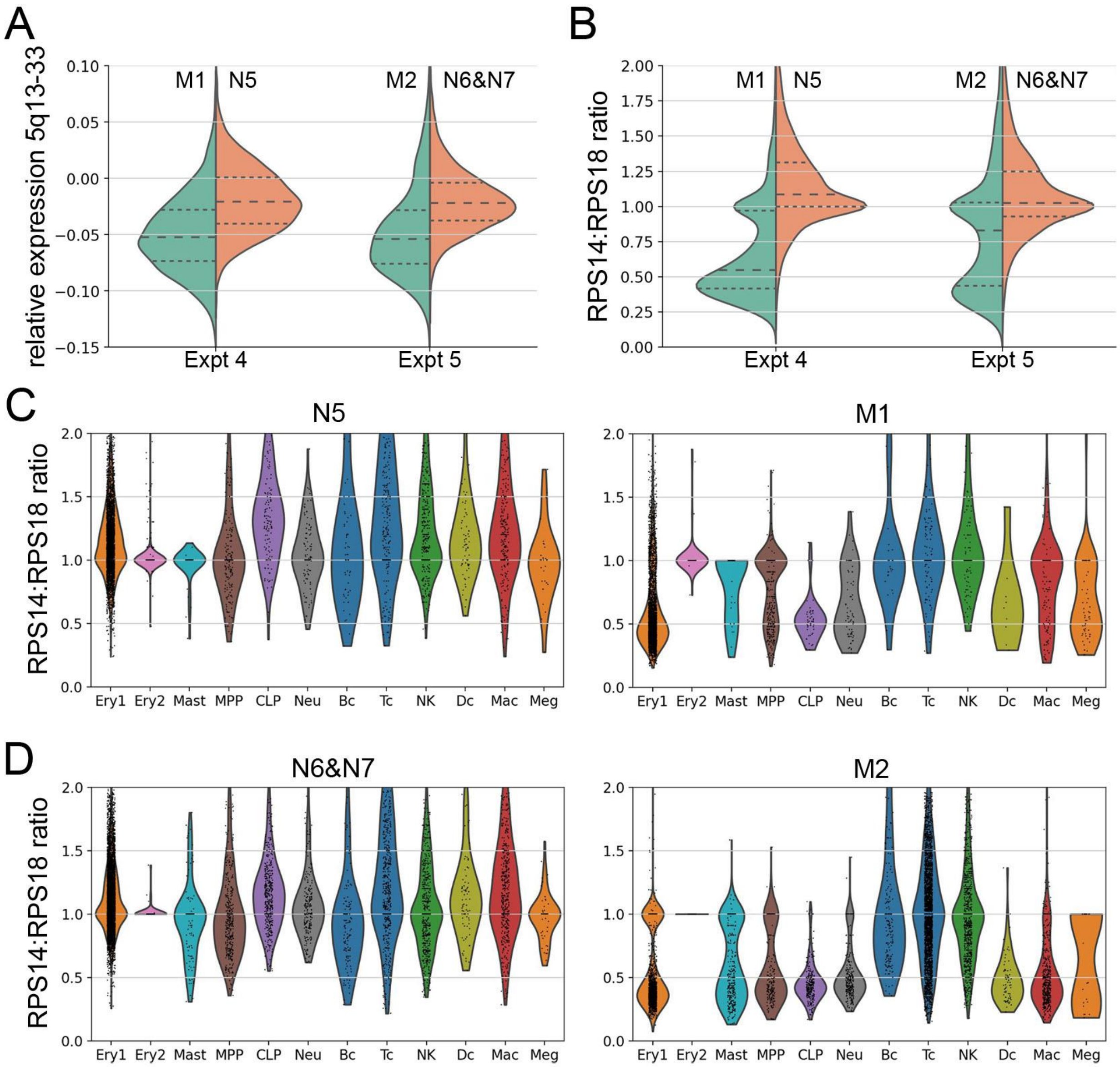


**Supplemental Figure 7. Deconvolution analysis of the del(5q) MDS sample identifies cells with the 5q13-33 deletion.** (**A**) Relative expression levels of genes in the 5q13-33 deletion interval according to the methods of Patel *et al.*[56] for the del(5q) MDS patients (M1 & M2) and normal individuals (N5, N6 & N7) show a significant reduction in gene expression in the del(5q) MDS patient cells. (**B**) The ratio of gene expression of RPS14 to RPS18 (Ch6p21.32) also shows significant reduction in RPS14 expression in cells from the del(5q) MDS patients, however it provides better separation between the $5q^+$ and $5q^-$ cells because high levels of RP expression is not diluted by low expression of other genes in the deletion interval. Analysis of haploinsufficient RPS14 expression in all marrow lineages from experiments 4 (**C**) and 5 (**D**) shows that surviving T cells, B cells, and NK cells (mostly ex vivo cells) in the del(5q) MDS patient have mostly intact 5q (normal RPS14 expression) while the other cell types have both $5q^+$ and $5q^-$ cells with little selective expansion. Because cells in the Ery2 cell population (late stage erythroblasts) are shutting down expression of all genes, including ribosomal protein genes, the near normal expression levels detected in these cells cannot be used to identify haploinsufficient cells accurately.
